# A recessive *PRDM13* mutation results in congenital hypogonadotropic hypogonadism and cerebellar hypoplasia

**DOI:** 10.1101/2021.07.28.21260126

**Authors:** Danielle E. Whittaker, Roberto Oleari, Louise C. Gregory, Polona Le Quesne-Stabej, Hywel J. Williams, GOSgene, John G. Torpiano, Nancy Formosa, Mario J. Cachia, Daniel Field, Antonella Lettieri, Louise Ocaka, Alyssa J.J. Paganoni, Sakina Rajabali, Kimberley L. Riegman, Lisa Benedetta De Martini, Taro Chaya, Iain C.A.F. Robinson, Takahisa Furukawa, Anna Cariboni, M. Albert Basson, Mehul T. Dattani

**Author notes:** Corresponding authors. Correspondence to: Professor Mehul Dattani, Section of Molecular Basis of Rare Disease, Genetics and Genomic Medicine Research & Teaching Department, UCL GOS Institute of Child Health, London, UK., Albert Basson, Anna Cariboni. Department of Molecular Medicine and Pathology, University of Auckland, Auckland, New Zealand (Polona Le Quesne-Stabej). Roche Pharma Research and Early Development (pRED), Neuroscience and Rare Diseases Discovery and Translational Area, Roche Innovation Center, F. Hoffmann-La Roche Ltd, Basel, Switzerland. Joint first authors (Danielle E. Whittaker, Roberto Oleari, Louise C. Gregory).

## Abstract

PRDM13 (PR Domain containing 13) is a putative chromatin modifier and transcriptional regulator that functions downstream of the transcription factor PTF1A, which in turn controls GABAergic fate in the spinal cord and neuronal development in the hypothalamus. Here, we report a novel, recessive syndrome associated with *PRDM13* mutation. Patients exhibited intellectual disability, ataxia with cerebellar hypoplasia, scoliosis and delayed puberty with congenital hypogonadotropic hypogonadism (CHH). Expression studies revealed *Prdm13/PRDM13* transcripts in the developing hypothalamus and cerebellum in mouse and human. We investigated the development of hypothalamic neurons and the cerebellum in mice homozygous for a *Prdm13* mutant allele.

A significant reduction in the number of Kisspeptin (Kiss1) neurons in the hypothalamus and PAX2+ progenitors emerging from the cerebellar ventricular zone were observed. The latter was accompanied by ectopic expression of the glutamatergic lineage marker TLX3. Phenotypically, mice lacking PRDM13 displayed cerebellar hypoplasia, normal gonadal structure, but delayed pubertal onset. Together, these findings identify PRDM13 as a critical regulator of GABAergic cell fate in the cerebellum and of kisspeptin neuron development in the hypothalamus, providing a mechanistic explanation for the co-occurrence of CHH and cerebellar hypoplasia in this syndrome. To our knowledge, this is the first evidence linking disrupted PRDM13-mediated regulation of Kiss1 neurons to CHH in humans.

## Introduction

Congenital hypogonadotropic hypogonadism (CHH) is a rare genetic disorder caused by defective development or functioning of hypothalamic gonadotrophin-releasing hormone (GnRH) secreting neurons, leading to deficiency of GnRH, the master hormone of the reproductive axis (1). During embryogenesis, GnRH neurons originating in the nasal placode migrate into the brain (2, 3) and innervate the median eminence to release GnRH. GnRH neurons receive several excitatory and inhibitory inputs from other hypothalamic neurons, such as Kisspeptin (Kiss1) (4, 5) which mainly promotes GnRH secretion, and Gonadotropin-Inhibitory Hormone (GnIH) neurons (6), which instead suppress GnRH neuronal activity. The interaction of their signalling pathways is expected to fine-tune GnRH neuronal activity, and consequently, pulsatile gonadotropin secretion from the pituitary gland (6). Disruption of some of these neuronal networks can lead to CHH (1, 7-9).

CHH is clinically characterized by complete or partial absence of puberty, and impaired or absent fertility. Additional clinical signs can be present in multi-syndromic forms of CHH (8), including ataxia, which is thought to be due to cerebellar hypoplasia (10-17). Early embryonic defects in cerebellar patterning have been linked to hypoplasia of the vermis, the medial portion of the cerebellum (18, 19), while alterations in progenitor cells tend to affect all regions of the cerebellum (20-22). Cerebellar neurons derive from two germinal zones, the rhombic lip and the ventricular zone. Glutamatergic neurons are specified in the rhombic lip, while GABAergic neurons originate in the ventricular zone. The basic-helix-loop-helix transcription factors ATOH1 and PTF1A contribute to the spatial segregation of these germinal zones (20, 22-24). While ATOH1 deletion leads to failure of glutamatergic progenitors to expand (22), PTF1A deficiency results in the mis-specification of GABAergic ventricular zone progenitors and aberrant expression of glutamatergic fate markers (21, 25).

The concurrence of ataxia, cerebellar atrophy or hypoplasia and specific endocrine disorders such as CHH has been previously reported (14-17). Cerebellar atrophy is present in rare conditions, such as Gordon Holmes (GHS), Oliver McFarlane and Boucher-Neuhauser syndromes, which appear to be caused by neurodegeneration. This far, gene mutations that can disrupt both cerebellar and hypothalamic development to cause cerebellar hypoplasia and CHH have not been reported.

*PRDM13* belongs to the PRDM family of transcriptional regulators, defined by a PR (positive regulatory) domain and a variable number of zinc finger domains (26, 27). PRDM factors modulate transcriptional activity by acting either as direct histone methyltransferases via catalytic activity of their PR domains (28, 29), or by recruiting other histone modifying enzymes to chromatin (30-33). They are involved in many developmental processes that drive and maintain cell state transitions or that modify the activity of signaling pathways (34). PRDM13 functions as an essential GABAergic cell fate determinant in the spinal cord and the retina (35-37). In the spinal cord, PRDM13 functions downstream of PTF1A to promote GABAergic and suppress glutamatergic fate (37, 38).

PTF1A is a critical ventricular zone specification factor during early cerebellum development, where it is both necessary and sufficient for the development of GABAergic neuronal populations (Purkinje cells and interneurons) (20, 21). Recessive mutations of PTF1A are associated with pancreatic and cerebellar agenesis/hypoplasia (39). Further, PTF1A was recently found to be required for the development of the hypothalamus in the mouse, where its forebrain-specific deletion leads to disruption of the Kiss1 neuronal system, hypogonadism and altered sexual behaviours, via a mechanism that does not involve a GABAergic/glutamatergic imbalance (40). These findings suggest the possibility that *PTF1A* or *PRDM13* mutations may be responsible for some congenital disorders characterized by both cerebellar hypoplasia and CHH.

Here, we report a novel, recessive syndrome associated with a *PRDM13* mutation in unrelated patients exhibiting intellectual disability, ataxia with cerebellar hypoplasia, scoliosis and delayed puberty with CHH. By combining exome sequencing on human patients with phenotypic analysis of *Prdm13*-deficient mice, we identified critical neurodevelopmental functions for PRDM13 that underlie these reproductive and cerebellar phenotypes. Our results are consistent with a conserved PTF1A-PRDM13 regulatory axis controlling cell fate specification in several neurogenic niches in the developing brain, including the hypothalamus and cerebellum.

## Results

### A novel recessive syndrome with PRDM13 mutation

We identified three patients (2 male, 1 female) from two unrelated families presenting with similar clinical features. Patients presented with delayed motor development, ataxia, scoliosis, intellectual disability and delayed sexual development (Supplemental Table 1). All patients required corrective surgery for scoliosis and presented with either generalised hypotonia with hyporeflexia (Patients 1 and 2) or hypertonia with hyperreflexia (Patient 3). Further details outlining the neurological findings are shown in Supplemental Tables 1-2. Patients 1 and 3 also had cerebellar hypoplasia on neuroimaging (Figure 1B-D).

**Figure 1.**
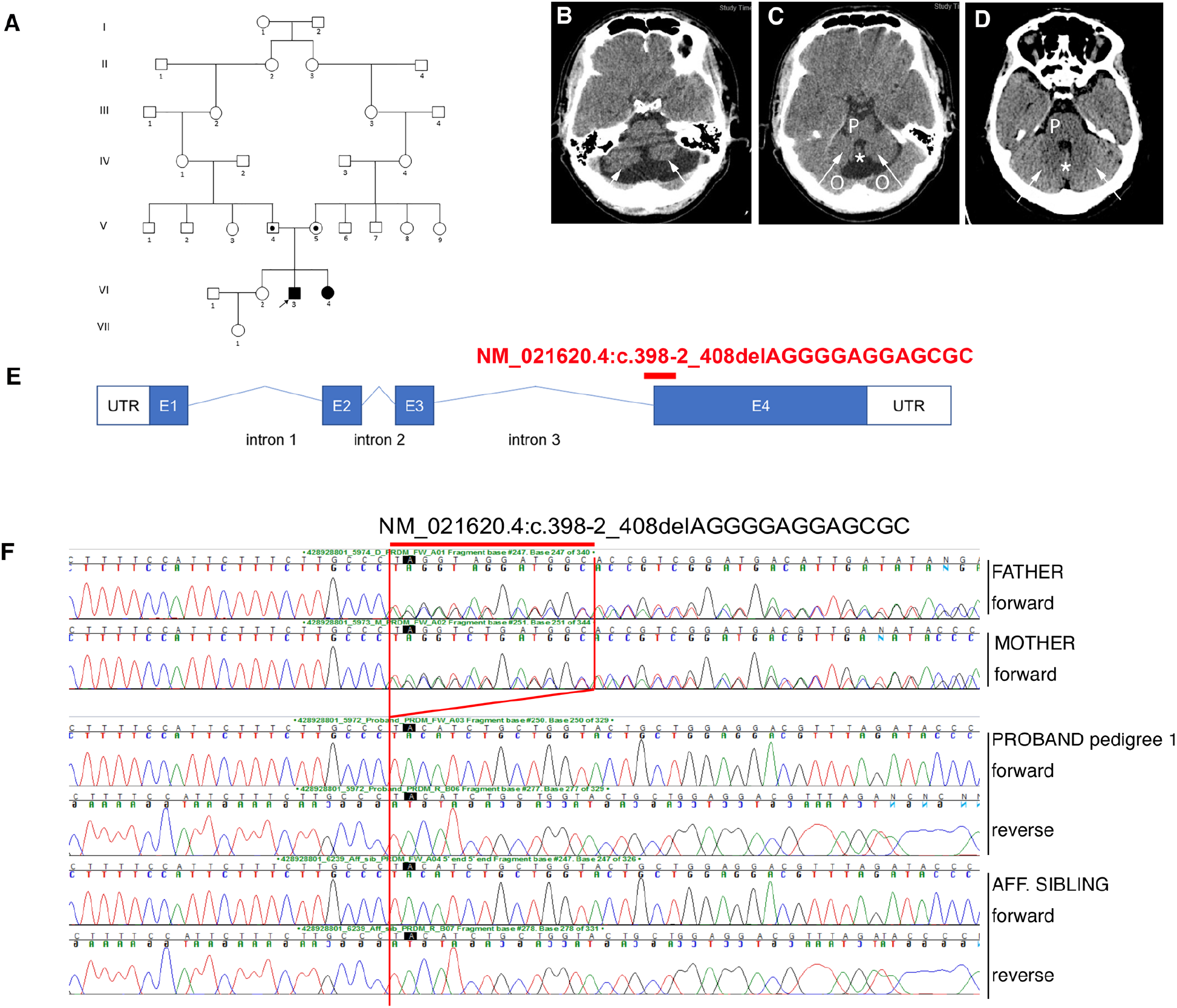
Exome sequencing identifies a PRDM13 mutation in three patients from 2 ethnically-matchedpedigrees. (**B-D**) (B-C) Axial slices on CT scan of the brain showing cerebellar hypoplasia in patient 1 compared with a normal CT scan (D). Arrows in (B-C) demonstrate hypoplastic cerebellar lobes as compared with those in the control scan (D). The pons (P) is hypoplastic compared with the scan in (D), as is the cerebellar vermis (asterisk, *). Partial voluming from the occipital lobes above the tentorium is seen (O). (D) CT scan showing normal cerebellar lobes (arrows), a normal cerebellar vermis (asterisk, *), and a normal pons (P). This normal scan was obtained courtesy of David Cuete, from Radiopaedia. (**E**) Diagram of *PRDM13* transcript (NM_021620.4) showing the deletion found in patients at intron 3-exon 4 border, which is predicted to affect splicing and to form a truncated PRDM13 protein.

CHH was diagnosed based on a combination of clinical and biochemical data. Patient 1 had bilateral undescended testes and underwent bilateral orchidopexies, which was repeated on the left between 5-10 years of age. CT scans of the brain revealed hypoplasia of the cerebellar hemispheres and vermis (Figure 1B). Progression of scoliosis necessitated a 2-stage surgical fixation of the spine between 5-10 years of age. He was referred to the Paediatric Endocrine Clinic at between 10-15 years with delayed puberty. Pubertal staging at this age was G1 P1 A1 -/03 mL A GnRH test revealed a peak LH of 2.3 IU/L, with an FSH of 4.4 IU/L. A 3-day hCG test revealed no change in the testosterone concentration after 3 hCG injections (peak testosterone of 2.2 nmol/L), and was therefore suboptimal and consistent with CHH. We have previously reported that a peak LH to GnRH stimulation of <2.8 IU/L, peak 3-day testosterone cut-off of <1.04 µg/L (3.6 nmol/L), and a peak 3-week testosterone cut-off of <2.75 µg/L (9.5nmol/L)] gave a sensitivity of 88% and a specificity of 100% for the diagnosis of CHH (41). Pubertal induction was commenced with testosterone supplementation at the age of 14.5 years.

Patient 2 was noted to have generalised hypotonia and hyporeflexia, as well as delayed gross motor development. A progressive right-sided thoraco-lumbar scoliosis was first noted at 0-5 years of age. All neurological investigations, including metabolic screen, EMG, and brain MRI were reported as normal. She was first seen in the Paediatric Endocrinology Clinic between 10-15 years of age, with a similar neurological condition to relative (Patient 1). She had not entered spontaneous puberty by the age of 12.5 years, with low basal gonadotrophins (basal LH <0.1 U/L, FSH 0.8 IU/L), and with a peak LH of 2.6 IU/L and an FSH of 6.4 IU/L on GnRH testing. Few data are available for cut-off values for the diagnosis of CHH in females, but these gonadotrophin responses would be considered to be sub-optimal in a 12.5 year old girl with no signs of puberty, as would the undetectable basal LH concentration. Pubertal induction was commenced with oestrogen supplementation.

Patient 3 presented with global developmental delay, generalised hypertonia and hyperreflexia between 0-5 years of age. A CT brain scan revealed cerebellar hypoplasia, and he had a broad-based gait. He needed corrective surgery for strabismus as well as spinal surgery for progressive scoliosis between10-15 years of age, but became completely wheelchair-dependent by the age shortly after surgery. He was referred to the Paediatric Endocrine Clinic at the age of 10-15 years with a micropenis. On pubertal staging at this age, his stretched penile length was 4 cm (less than P10) and both testes were impalpable. Basal gonadotrophins were low (LH <0.1 IU/L, FSH 0.4 IU/L). A GnRH test performed at 10-15 years revealed a peak LH of 0.9 IU/L with an FSH of 3.1 IU/L. The peak testosterone was sub-optimal at 2.3 nmol/L after a 3 day hCG test, with an excellent peak of 30.7 nmol/L after 3 weeks of HCG. The sub-optimal LH together with sub-optimal response to 3 days of HCG and the micropenis with undescended testes support a diagnosis of CHH. Following these tests, the left testis descended into the scrotum (2 mL volume), but the right testis remained impalpable. Pubertal induction with testosterone enantate was commenced at low dose by intramuscular injection, and the dose increased gradually over the following 2 years. Bilateral orchidopexies were performed at 10-15 years of age. Over time, he experienced penile growth, but both testes remained 2 mL in volume.

### Whole exome sequencing of patients 1 and 2 identified a novel 13bp deletion in PRDM13

The homozygous *PRDM13* deletion: NC_000006.11:g. 100060906_ 100060918del (c.398-3_407delCAGGGGAGGAGCG), located at chr6:100060906 (GRCh37), spans the intron3/exon4 boundary (Figure 1E). This pathogenic variant is predicted to affect splicing with premature truncation of PRDM13 resulting in the loss of all four Zn finger domains according to Mutation Taster software (42). This variant was confirmed by Sanger sequencing to be homozygous in patients 1 and 2, and heterozygous in the unaffected parents respectively. Given the phenotypic similarity of Patient 3 to 1 and 2, we opted to perform Sanger sequencing of *PRDM13* in patient 3. This confirmed the presence of the same homozygous 13bp deletion in this patient. His unaffected parents were also heterozygous for the deletion. This mutation was not present in control databases, including the gnomAD browser (https://gnomad.broadinstitute.org/) (∼123k samples). To determine if this mutation might be present in the same ethnically-matched population, forty-two control individuals were screened for this *PRDM13* variant in the same population, one of whom was a heterozygous carrier. To delineate the region harbouring the 13bp deletion in more detail, we performed genome-wide microarray analysis of the three patients and an unrelated heterozygous carrier from the same ethnic background. This revealed that the three patients shared an identical region of homozygosity, spanning approximately 1.6Mb that encompassed the 13bp deletion. The heterozygous carrier shared a smaller nested region of homozygosity of 0.2Mb also encompassing the 13bp deletion (Supplemental Table 3).

### A homozygous Prdm13 mouse model for investigating non-lethal phenotypes

Mice homozygous for *Prdm13* null mutations are perinatal lethal (37), suggesting that the *PRDM13* mutations in these patients are most likely partial loss-of-function. The only homozygous *Prdm13* mutations thus far reported that do not cause perinatal lethality in mice are those with frameshift mutations in exon 1 or deletion of exons 2 and 3 (35, 37). We therefore generated homozygous *Prdm13* mutants carrying targeted deletion of exons 2 and 3, encoding the majority of the PR domain (Supplemental Figure 1A, B). Quantitative reverse transcriptase PCR (qRT-PCR) confirmed the absence of exon 2 and 3 transcripts and the presence of exon 4-containing transcripts in *Prdm13*^*Δex2,3*^*/* ^*Δex2,3*^ mutant cerebella at E12.5 (Supplemental Figure 1C). Immunostaining of cerebellar tissue from these mice (Figure 5H,J) with antiserum raised against the C-terminal fragment of the protein (aa685-754) suggested the absence of this epitope. These *Prdm13*^*Δex2,3*^*/* ^*Δex2,3*^ mutants, referred to from here on as *Prdm13*^*-/-*^ mutants, survived to adulthood as previously reported (35). *Prdm13*^*-/-*^ mice did not exhibit any signs suggestive of a neurodegenerative disease associated with ageing (age range 8-12 months, n=9).

### Prdm13 loss does not affect the development of GnRH neurons

As patients carrying homozygous mutations of *PRDM13* display gonadotropin deficiency (Supplemental Table 1), we first analyzed the GnRH neuron system in *Prdm13*^*-/-*^ mutants. Expression of *prdm13* has been reported in the olfactory placode that gives rise to GnRH neurons in zebrafish (43). We detected *Prdm13* expression by RT-PCR both in the mouse nasal compartment and forebrain during developmental stages relevant to GnRH neuron migration (Figure 2A). Immunohistochemical detection of GnRH neurons in coronal sections at E14.5 revealed normal numbers of GnRH neurons in *Prdm13* mutants (GnRH neuron number mean ± SEM.: *Prdm13*^+/+^ 1341.33 ± 41.58 vs. *Prdm13*^-/-^ 1453.00 ± 37.03; P=0.1154, two-tailed unpaired Student’s *t* test, n=3 for each group). Because GnRH neurons at E14.5 can be observed in the nose, in the nasal-forebrain junction (nfj) and in the medial preoptic area (mpoa) of the brain, we next determined the relative number of neurons in these compartments and observed a similar number of GnRH-positive cells in the nose, in the nfj and in the mpoa of mutants compared to wild-type mice (Figure 2B-G; GnRH neuron number mean ± SEM: <u>nose</u>: *Prdm13*^+/+^ 406.33 ± 32.10 vs. *Prdm13*^−/–^ 431.33 ± 7.84, P=0.4914; <u>nfj:</u> *Prdm13*^+/+^ 378.67 ± 24.36 vs *Prdm13*^−/–^ 413.67 ± 27.63, P=0.3958; <u>mpoa</u>: *Prdm13*^+/+^ 556.33 ± 92.35 vs. *Prdm13*^−/–^ 608.00 ± 10.02, P=0.6077; two-tailed unpaired Student’s *t* test, n=3 for each group). Normal GnRH neuron development is reflected postnatally in a normal innervation of the median eminence, the region where GnRH neurons release the decapeptide GnRH into the portal blood vessels of the pituitary. A comparison of GnRH neuron projections to the median eminence in postnatal day (P) 22 *Prdm13*^−/–^ and *Prdm13*^+/+^ littermates found no differences between the two genotypes (Figure 2H,I). Altogether, these data suggest that *Prdm13*, although expressed during GnRH neuron development, does not play a prominent role in controlling their number or migration.

**Figure 2:**
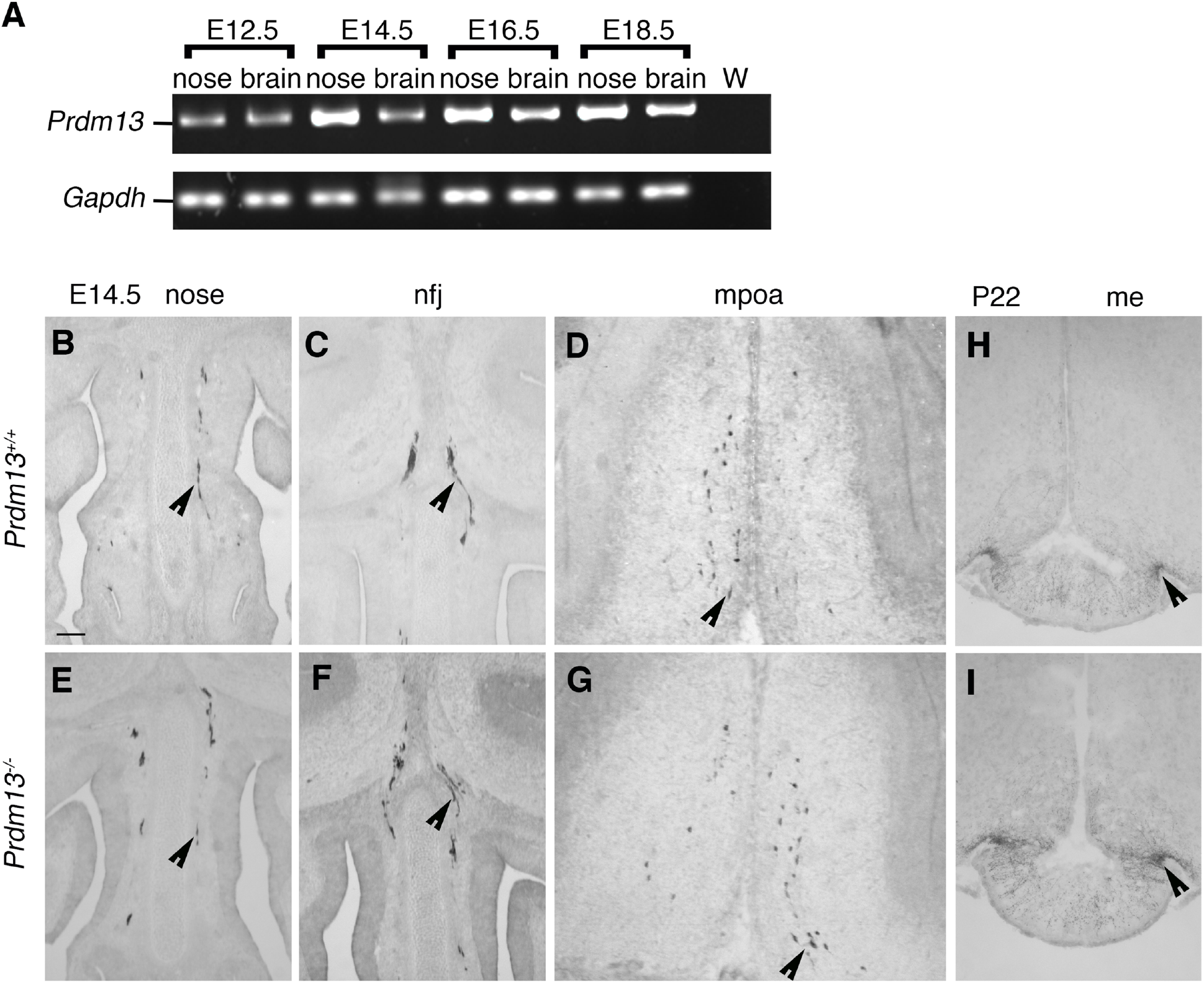
GnRH neuron specification and migration are not affected in *Prdm13* mutant mice. **(A)** RT-PCR analysis of *Prdm13* expression in mouse embryonic nose and brain tissues extracted from indicated embryonic stages of wild-type mouse embryos. *Gapdh* expression serves as positive control. **(B-G)** Coronal sections of *Prdm13*^*+/+*^ and *Prdm13*^*-/-*^ E14.5 heads, immunolabelled for GnRH to reveal GnRH neurons in the nasal compartment (B,E), in the NFJ (C,F) and in the MPOA (D,G). Black arrows indicate examples of neurons migrating in the nasal compartment (B-E), crossing the olfactory bulbs (C,F) and reaching the MPOA (D,G). **(H**,**I)** Coronal sections of *Prdm13*^*+/+*^ and *Prdm13*^*-/-*^ P22 brains immunolabelled for GnRH to reveal ME innervation by GnRH neuron neurites. Scale bars: 250 µm. W = water only control, no cDNA; nfj = nasal-forebrain junction; mpoa = medial preoptic area; me = median eminence.

### Prdm13 loss affects the development of Kiss1 neurons in the hypothalamus

Microarray data have shown that *Prdm13* expression is enriched in the adult dorso-medial (DM) and, to a lesser extent, in the arcuate (Arc) nuclei (44) of the hypothalamus where GnIH and Kiss1 neurons are found, respectively. Thus, we asked if GnRH deficiency might be indirectly caused by the lack or malfunctioning of hypothalamic neurons that regulate GnRH secretion. First, we visualized *Prdm13* expression in the adult brain by *in situ* hybridization, which confirmed *Prdm13* expression in the Arc and DM nuclei of the adult hypothalamus (Figure 3A).

**Figure 3:**
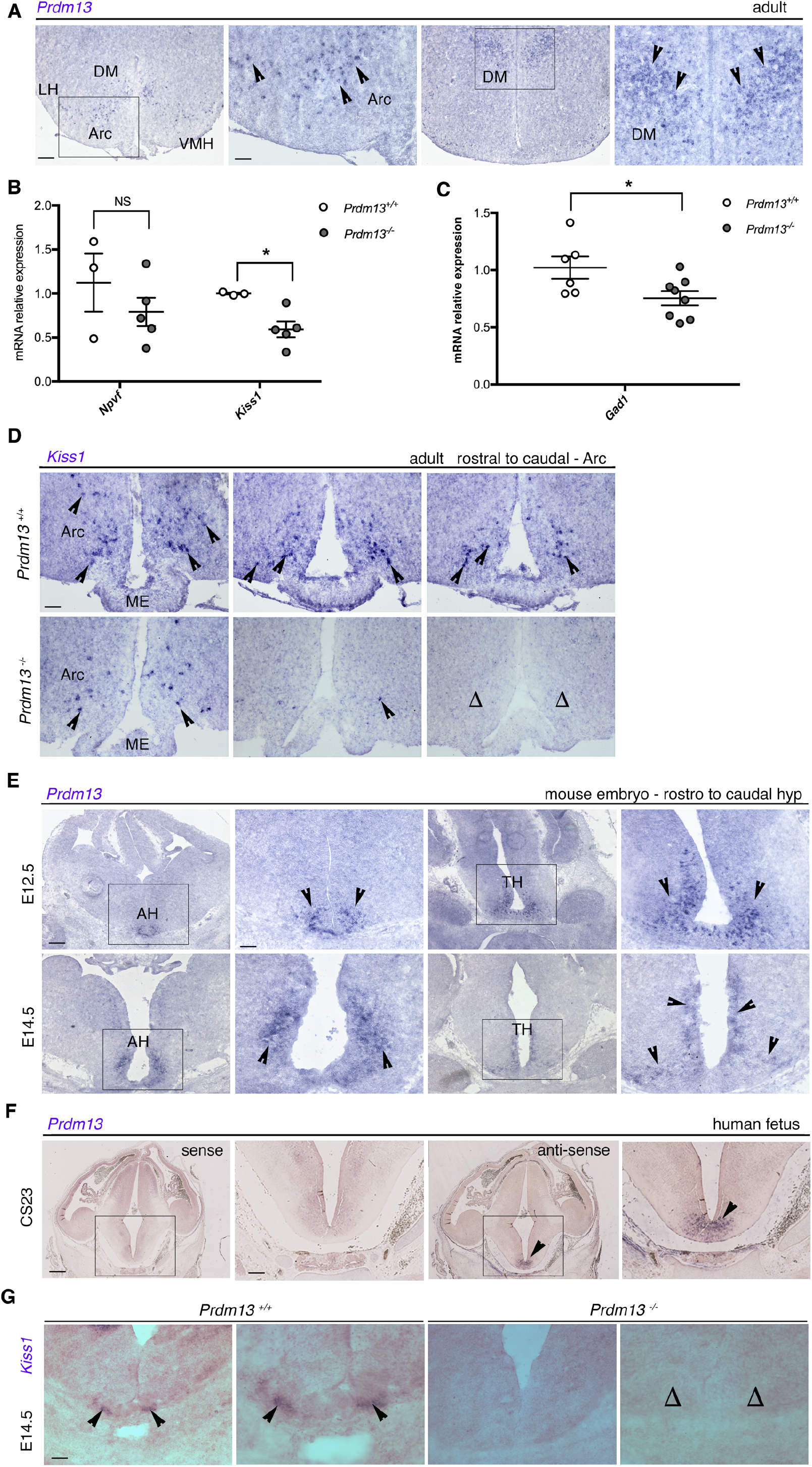
*Prdm13* loss affects *Kiss1* expression and *Kiss1* neuron development. **(A)** *In situ* hybridization experiments on coronal adult wild-type male mice brain sections to detect the expression of *Prdm13* in the Arc and DM nuclei of the hypothalamus. High magnification of the squared areas are shown next to each panel. **(B)** qRT-PCR analysis for *Npvf* and *Kiss1* transcripts in the hypothalamus of *Prdm13*^*+/+*^ and *Prdm13*^*-/-*^ male mice. ΔΔCq were calculated relative to control samples using quantification cycle (Cq) threshold values that were normalised to the housekeeping gene, *Gapdh*. Note the significant decrease of *Kiss1* levels in mutants. **(C)** qRT-PCR analysis for *Gad1* transcripts in the hypothalamus of *Prdm13*^*+/+*^ and *Prdm13*^*-/-*^ from both sexes. ΔΔCq were calculated relative to control samples using Cq threshold values that were normalised to the housekeeping gene, *Gapdh*. Note the significant decrease of *Gad1* levels in mutants. **(D)** *In situ* hybridization experiments on sections taken at the level of the ARC nucleus from *Prdm13*^*+/+*^ and *Prdm13*^*-/-*^ male mice for *Kiss1* transcripts. Note the reduction in *Kiss1* expression in the mutants, compared to wild-type controls. **(E-F)** *In situ* hybridization on coronal sections to detect *Prdm13/PRDM13* expression in the developing mouse hypothalamus at E12.5 and E14.5 (E), and in the developing human hypothalamus at CS23 (F), mRNA transcripts are indicated by the arrowheads. The sense probe showed negative staining (the first two images from the left in (F)). The areas in the marked rectangles are shown in high magnification to the right of the respective image. **(G)** *In situ* hybridization on coronal sections from E14.5 mouse embryo to detect *Kiss1* expression in *Prdm13*^*+/+*^ and *Prdm13*^*-/-*^. Black arrowheads indicate examples of Kiss1-expressing cells; note the absence of Kiss1 neurons in mutants (Δ). *P<0.05, Student’s t test. Scale bars: 500 μm (A-E-F low magnification), 250 μm (A-E-F high magnification, D, G). LH = lateral hypothalamus; DM = dorsomedial nucleus; Arc = arcuate nucleus; VMH = ventromedial hypothalamus; ME = median eminence; AH = anterior hypothalamus; TH = tuberal hypothalamus.

We then assessed if *Prdm13* could regulate *Npvf* (which encodes GnIH) and *Kiss1* expression. First, we dissected the hypothalamus from adult male *Prdm13*^-/-^ and *Prdm13*^+/+^ mice and quantified *Npvf* and *Kiss1* expression by qRT-PCR. This analysis showed a significant reduction of *Kiss1* mRNA levels in *Prdm13*^-/-^ mice compared to *Prdm13*^+/+^ mice. *Npvf* expression appeared slightly reduced but this difference was not significant (Figure 3B). Because PRDM13 is known to control GABAergic neuronal cell fate in several brain areas (35-37), and GABAergic neurotransmission plays important roles to sustain fertility (45), we also analyzed *Gad1* expression and found a significant decrease of *Gad1* levels in *Prdm13*^-/-^ mice compared to *Prdm13*^+/+^ mice (Figure 3C). However, the expression of typical GABAergic hypothalamic neuronal markers such as *POMC* and *NPY/AgRP* (46), was unchanged between *Prdm13*^-/-^ and *Prdm13*^+/+^ mice (Supplemental Figure 2A), suggesting that PRDM13 is required for a specific subset of hypothalamic neurons.

Next, we confirmed the reduction of *Kiss1* levels by visualizing *Kiss1*-expressing neurons in adult male mouse brains by *in situ* hybridization. This analysis, although qualitative, clearly revealed the presence of fewer *Kiss1*-expressing cells in the Arc nucleus of *Prdm13*^-/-^ mice compared to *Prdm13*^+/+^ controls (Figure 3D). Kiss1 neurons can be found in the anteroventral periventricular (AVPV) nucleus of the hypothalamus as well (47), where they contribute to GnRH secretion in both sexes, as well as increasing their activity during the GnRH/LH surge in females (48). We checked *Prdm13* expression in this region, but did not detect *Prdm13* transcripts in the AVPV nucleus of P15 and adult brains (Supplemental Figure 2B). Accordingly, the position and number of *Kiss1*^+^ neurons was unchanged in this region in *Prdm13*^*-/-*^ mutants (Supplemental Figure 2C-D; *Kiss1*^+^ neuron number mean ± SEM: *Prdm13*^+/+^ 238.3 ± 38.62 vs. *Prdm13*^−/–^ 218.3 ± 8.67; P=0.6399, two-tailed unpaired Student’s *t* test, n=3 for each group). Consistent with an unchanged number of AVPV *Kiss1*^+^ neurons, which are thought to be more dense in females (48), the overall hypothalamic *Kiss1* expression was reduced, however not significantly, in *Prdm13*^*-/-*^ females compared to *Prdm13*^+/+^ controls (Supplemental Figure 2E).

To determine if PRDM13 had a role in the development of Kiss1 neurons, we further examined the expression of *Prdm13/PRDM13* in the developing mouse and human hypothalamus. *Prdm13* is expressed in the preoptic and tuberalis nuclei of the developing mouse hypothalamus (Figure 3E). We also detected prominent *PRDM13* expression in the developing hypothalamus of a human fetus at Carnagie stage (CS)23 (Figure 3F). The first *Kiss1-*expressing cells can be detected as early as E13.5 near the third ventricle of the embryonic mouse brain (49), with numbers increasing as development proceeds. Thus, we visualized Kiss1 neurons in the brains of E14.5 mice by *in situ* hybridization and found that the number of these cells in the presumptive Arc nucleus of *Prdm13*^-/-^ embryos was reduced substantially compared with wild-type littermates (Figure 3G; *Kiss1*^+^ neuron number mean ± SEM.: *Prdm13*^+/+^ 79.33 ± 8.37 vs. *Prdm13*^−/–^ 0.00; *** P=0.0007, two-tailed unpaired Student’s *t* test, n=3 for each group). To get insights into the possible mechanism through which PRDM13 affects the number of Kiss1 neurons, we compared cell proliferation and apoptosis in the developing hypothalamus between E14.5 *Prdm13*^+/+^ and *Prdm13*^−/–^ embryos, via immunodetection of phosphohistone 3B (PH3B) and cleaved caspase-3 (CC3) positive cells. As displayed in Supplemental Figure 3, we observed no differences in the number of PH3B^+^ and CC3^+^ cells between the genotypes, suggesting a differentiation defect rather than reduced precursor proliferation or increased cell death.

Altogether, these findings demonstrate that PRDM13 is required for the development of *Kiss1*-expressing neurons in the Arc nucleus of the hypothalamus. As Kiss1 promotes GnRH secretion, a deficiency in Kiss1 neurons in patients with homozygous *PRDM13* mutation may underlie the human CHH phenotype.

### Prdm13 null mice display normal gonadal structure but delayed vaginal opening

The loss of kisspeptin can result in abnormal sexual maturation, with significant phenotypic variability (50). An examination of testis size and morphology of adult post-pubertal *Prdm13*^-/-^ mice did not detect significant differences in gonadal size between P60 *Prdm13*^-/-^ mice and wild-type littermates (Figure 4A-A’). Furthermore, *Prdm13*^-/-^ mice displayed normal spermatogenesis, as assessed by H&E staining and immunodetection of the sertoli marker SOX9, which, together with SOX8, are essential for testis formation and spermatogenesis (51) (Figure 4B-C’). We also analysed the gonads of adult *Prdm13*^-/-^ females. The ovaries visually appeared slightly smaller in size, with a normal histology (Figure 4D-E’), but displayed a non-significant reduction in their weights (Figure 4F; weight mean±SEM = *Prdm13*^+/+^ 0.0098 ± 0.001 vs. *Prdm13*^−/–^ 0.0077 ± 0.00005; P=0.15, two-tailed unpaired Student’s *t* test, n=3 for each group). A significant delay in the timing of vaginal opening was observed in *Prdm13*^-/-^ females, compared to *Prdm13*^*+/+*^ controls (Figure 4F; vaginal opening day mean±SEM = *Prdm13*^+/+^ 27.55 ± 0.87 vs. *Prdm13*^−/–^ 33.91 ± 0.84; *** P<0.0001, two-tailed unpaired Student’s *t* test, n=11 for each group). These observations suggest that the reduction in *Kiss1* neurons in *Prdm13*^-/-^ mice was not sufficient to cause a gross gonadal phenotype, but induced a delay in pubertal onset.

**Figure 4.**
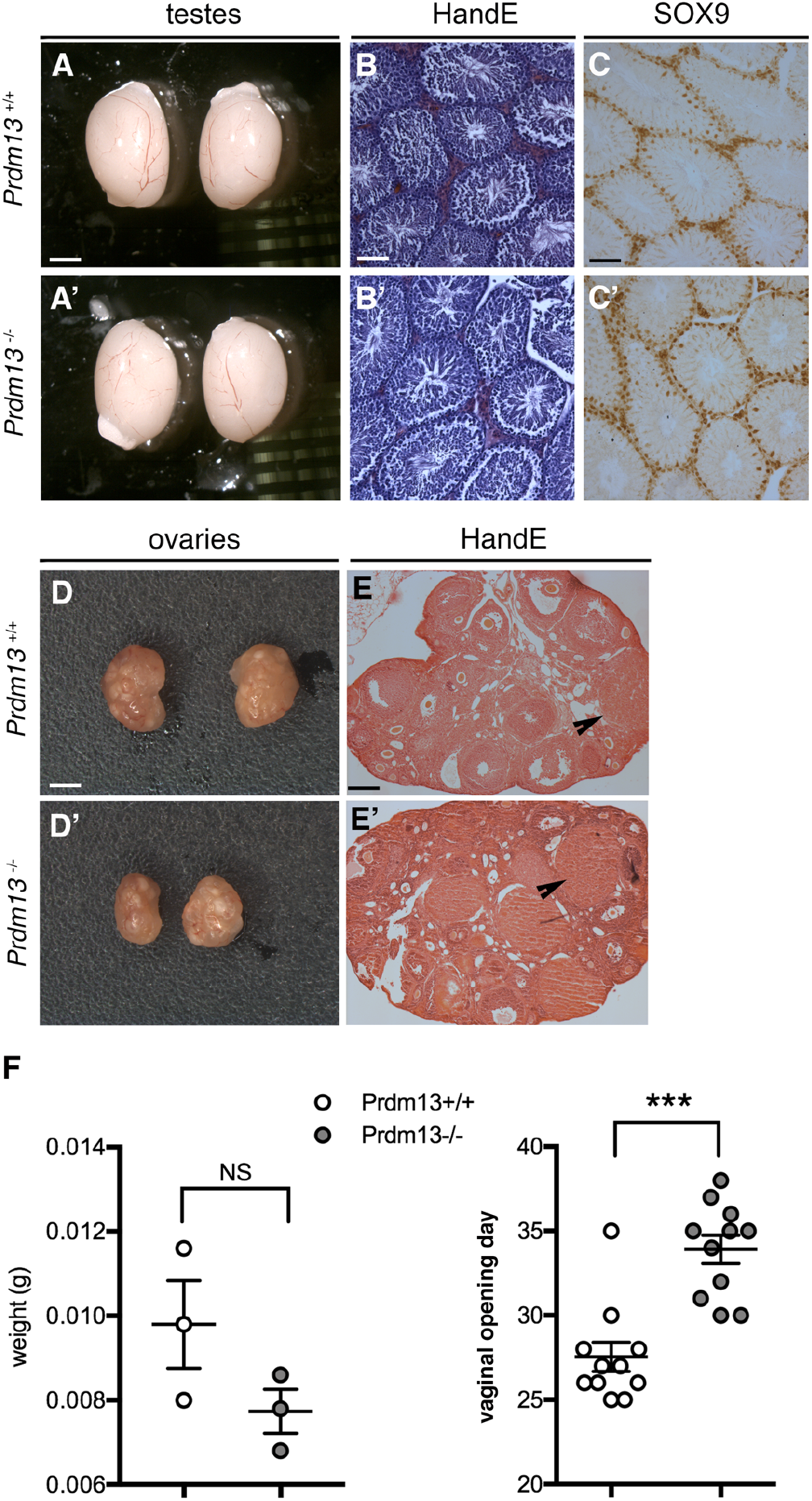
*Prdm13* deficient mice display normal gonadal structure, but delayed vaginal opening. **(A)** Microphotograph of testes from *Prdm13*^*+/+*^ and *Prdm13*^*-/-*^ mice respectively; no differences were observed in their size. **(B-C)** Hematoxylin and eosin (H&E)-stained (B) and SOX9-immunostained (C, Sertoli cells marker) testes representative sections from adult male mice of indicated genotypes. There was no difference, and normal spermatogenesis were observed, between wild-type and mutant mice. **(D)** Microphotograph of ovary pairs of the indicated genotypes. **(E)** H&E-stained ovary representative sections from mice of indicated genotypes. There was no difference in the number of corpora lutea observed between wild-type and mutant mice. **(F)** Weight of ovaries (left graph) and age at the time of the vaginal opening (VO) (right graph) from adult female mice of the indicated genotypes. Note the significant delay in the VO of *Prdm13*^*-/-*^ female mice. ***P<0.001. Scale bars: 1.5mm (A,E), 250μm (B,C), 500μm (F).

### Prdm13 expression in the human and mouse cerebellar ventricular zone

To understand how *PRDM13* deficiency may lead to cerebellar hypoplasia, we first examined *PRDM13* expression in human embryonic cerebellar tissue at CS23 and at equivalent stages in the mouse. *PRDM13* was expressed in the human cerebellar primordium (Figure 5A,B). Similarly, *Prdm13* expression was observed in the mouse embryonic cerebellum, with *Prdm13* transcripts detected prominently in the ventricular zone of both the cerebellar vermis and hemispheres (Figure 5C-F). Immunostaining with antiserum raised against a C-terminal fragment of PRDM13 (35) similarly revealed expression in cerebellar anlage at E12.5 (Figure 5G). At postnatal stages, PRDM13 localised to individual cells in the prospective white matter (Figure 5I,I’). The specificity of immunostaining was confirmed by the absence of staining in the cerebellum of *Prdm13*^-/-^ mice (Figure 5H,J,J’).

**Figure 5.**
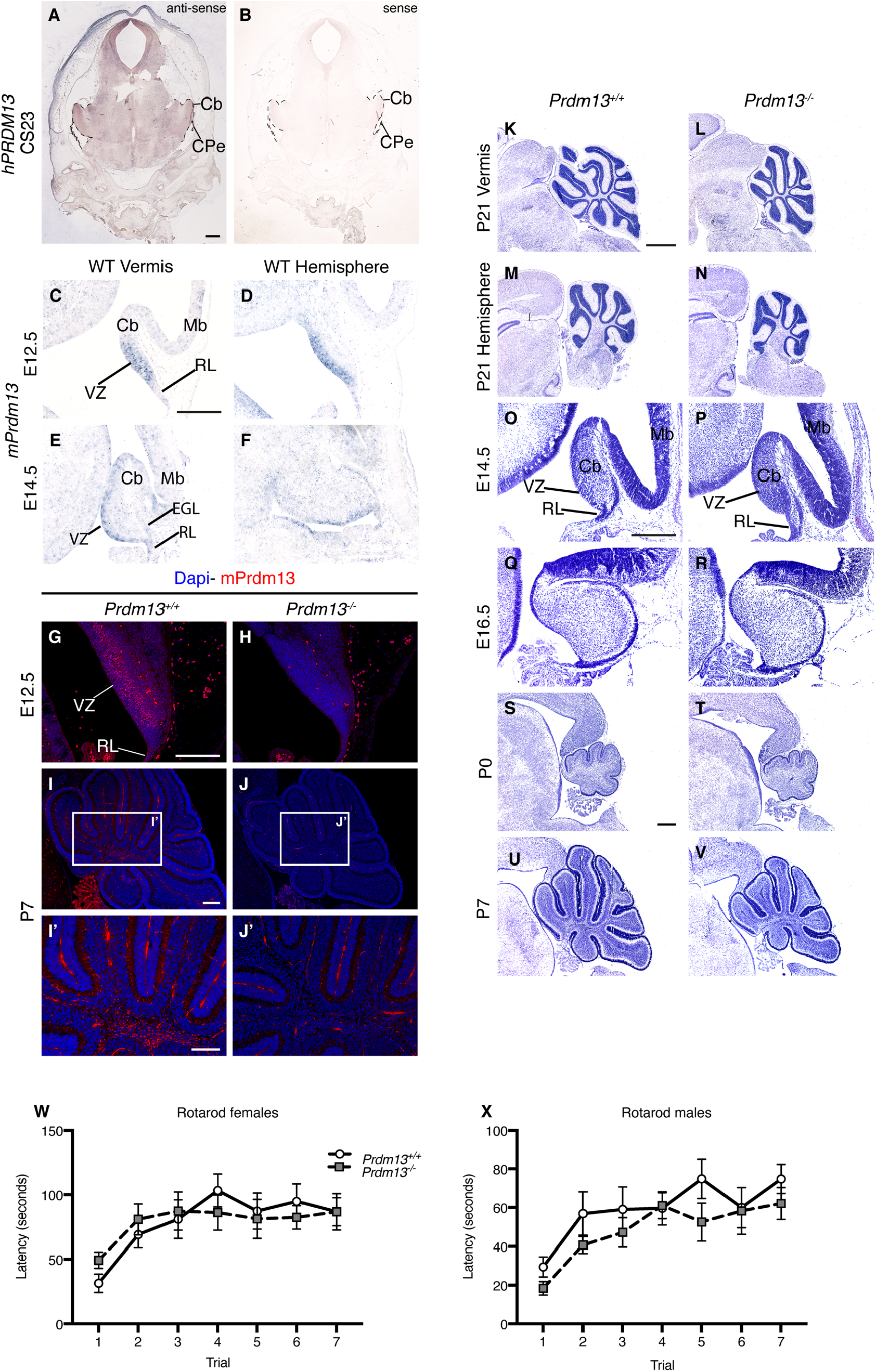
(A) *PRDM13* has a conserved role in regulating cerebellar growth during development. *In situ* hybridisation for human *PRDM13* (*hPRDM13*) transcripts (brown) in a coronal section through the cerebellum at Carnegie stage (CS) 23. **(B)** The *hPRDM13* sense control showed no *PRDM13* transcript staining throughout the developing cerebellum. **(C-F)** *In situ* hybridisation for *Prdm13* exon 4 transcripts (blue) in sagittal sections through the vermis and hemisphere of *Prdm13*^*+/+*^ cerebella, at stages indicated. **(G-J)** Immunohistochemistry of sagittal sections of *Prdm13*^*+/+*^ and *Prdm13*^*-/-*^ cerebella using PRDM13 antiserum at indicated stages. High power images are shown at postnatal day 7 (P7). Note that *Prdm13* transcripts (C-F) and protein (G) are predominantly restricted to the ventricular zone at mid-embryonic stages and to the cerebellar white matter postnatally (I). **(K-N)** Cresyl violet stained sagittal sections through the cerebellum of P21 *Prdm13*^*+/+*^ and *Prdm13*^*-/-*^ mice, anterior to the left. Note hypoplasia of the cerebellar vermis and hemispheres in *Prdm13*^*-/-*^ mice. **(O-V)** Time course through cerebellar development using Cresyl violet stained sagittal sections of the cerebellar vermis of *Prdm13*^*+/+*^ and *Prdm13*^*-/-*^ mice. Note that cerebellar hypoplasia is clearly evident in postnatal stages in *Prdm13*^*-/-*^ mice. **(W**,**X)** The mean latency of female (W) and male (X) mice to remain on the Rotarod over the course of seven trials (n=10 of each sex and genotype). Note that there is no difference between genotypes in female or male mice (2-way repeated measures ANOVA for genotype and trial). VZ = ventricular zone, RL = rhombic lip, Mb = midbrain, EGL = external germinal layer, Cb = cerebellum, CPe = Choroid plexus. Scale bar = 600μm (A), 300μm (C,G,I, I’,O,S), 1mm (K).

### Prdm13 is required for normal postnatal cerebellar growth

Next, we asked if *Prdm13*^-/-^ mice presented with cerebellar hypoplasia. In common with human patients with homozygous *PRDM13* mutations, *Prdm13*^*-/-*^ mice exhibited hypoplasia of the cerebellar vermis and hemispheres at P21, when cerebellar development is largely complete (Figure 5K-N). To identify the onset of reduced cerebellar growth in *Prdm13* mutants, cerebellar development was analysed from E14.5 to P7 (Figure 5O-V; Supplemental Figure 4). Cresyl-violet stained sagittal sections were measured in early postnatal stages to obtain an estimate of cerebellar surface area. Cerebellar surface area was not significantly altered at birth in the cerebellar vermis (mean±SEM = 0.51±0.02mm^2^ in *Prdm13*^*+/+*^, 0.51±0.03mm^2^ in *Prdm13*^*-/-*^; P=0.9259, two-tailed unpaired Student’s *t* test, n=4 for each group) or cerebellar hemispheres (mean±SEM = 0.69±0.02mm^2^ in *Prdm13*^*+/+*^, 0.54±0.09mm^2^ in *Prdm13*^*-/-*^; P=0.1181, two-tailed unpaired Student’s *t* test, n=4 for *Prdm13*^*+/+*^ and n=3 for *Prdm13*^*-/-*^ mice). Analysis at later stages revealed that the onset of the phenotype differed between cerebellar regions. By P5, hypoplasia was evident in the vermis (mean±SEM = 1.71±0.02mm^2^ in *Prdm13*^*+/+*^, 1.27±0.04mm^2^ in *Prdm13*^*-/-*^; *** P=0.0007, two-tailed unpaired Student’s *t* test, n=3 for each group), whilst hemisphere size did not significantly differ between genotypes at the same stage (mean±SEM = 1.32±0.17mm^2^ in *Prdm13*^*+/+*^, 1.17±0.06mm^2^ in *Prdm13*^*-*^*/-*; P=0.4131, two-tailed unpaired Student’s *t* test, n=3 for each group) (Supplemental Figure 4M-P). By P7 cerebellar hypoplasia was, however, clearly evident in both the cerebellar vermis (mean±SEM= 2.63±0.09mm^2^ in *Prdm13*^*+/+*^, 1.95±0.07mm^2^ in *Prdm13*^*-/-*^; **P=0.0049, two-tailed unpaired Student’s *t* test, n=3 for each group) and hemispheres (mean±SEM = 2.40±0.18mm^2^ in *Prdm13*^*+/+*^, 1.80±0.09mm^2^ in *Prdm13*^*-/-*^; *P=0.0377, two-tailed unpaired Student’s *t* test, n=3 for each group) (Figure 5U,V; Supplemental Figure 4Q-T). To investigate the mechanism through which *Prdm13* deficiency leads to cerebellar hypoplasia, proliferation and cell death were assessed at E16.5, P0 and P5, prior to and at the onset of the phenotype. Proliferating PH3B+ and apoptotic CC3+ cells in the external germinal layer (EGL), where glutamatergic granule neuron progenitors expand, and the rest of the cerebellum were counted separately. Proliferation was not significantly altered in the EGL or non-EGL cells of the cerebellar vermis and hemispheres of *Prdm13* mutants at any stage (Supplemental Figure 5A-D). In contrast the number of apoptotic cells was significantly increased in non-EGL cells in both the vermis and hemispheres of *Prdm13*-deficient cerebella at P0 (Supplemental Figure 5F,H,J,J’,L,L’). Together, these data indicate that *Prdm13* is required for normal postnatal cerebellar development, and that reduced growth in *Prdm13*^*-/-*^ mutants occurs, at least in part, due to increased apoptosis of cells outside the EGL.

To determine if the cerebellar hypoplasia in these mutants is associated with motor coordination and learning deficits, we tested mice on an accelerating Rotarod. Adult *Prdm13*^*-/-*^ mice of both sexes exhibited normal motor coordination and motor learning in this test (Figure 5W,X). These findings suggest that the cerebellar changes in *Prdm13*^*-/-*^ mutants were not sufficient to cause overt motor deficits.

### Prdm13 regulates GABAergic cell fate determination in the developing cerebellum

As *Prdm13* expression was largely confined to cerebellar GABAergic neuron progenitor zones in the embryonic cerebellum (Figure 5A-F), the number and distribution of the two major GABAergic lineages (PAX2+ interneurons and LHX1/5+ Purkinje cell (PC) progenitors) were assessed. First, LHX1/5+ immunostaining was used to visualize PC progenitors that have left the ventricular zone at three key stages of development: E14.5, when cerebellar corticogenesis is initiated by formation of the Purkinje plate (PP), a transient structure comprising several layers of PCs; P0, just prior to PC monolayer formation; and P7, when the PC monolayer has been established. Analysis revealed normal PP formation in *Prdm13*^*-/-*^ mice (Supplemental Figure 6A-H) with no apparent alteration in PC distribution and organization at birth (Supplemental Figure 6I-P), or alteration in the PC monolayer postnatally (Supplemental Figure 6Q-T). These findings suggest that *Prdm13* is not required for PC formation, migration and organization. PC dendritogenesis is completed during the second and third postnatal weeks in mice. To investigate if *Prdm13* influences PC differentiation, dendritogenesis was analysed using molecular layer thickness as a measure of dendritic span (52). No significant difference was detected between genotypes. These findings suggested that PC differentiation was unaffected by *Prdm13* deficiency, but we cannot completely exclude subtle abnormalities at this stage (Supplemental Figure 6U-Y). Next, the PAX2+ population was visualized from the onset of their specification (53, 54). At E12.5, PAX2+ cells were present in a small cluster in the lateral cerebellum in the wild-type mice (Supplemental Figure 7A-D); however such cells were absent from *Prdm13*^*-/-*^ cerebella at the same stage (asterisks in Supplemental Figure 7E-H). Later, at E14.5, PAX2+ cells were present as a thin layer throughout the dorsal – ventral aspect of the ventricular zone along the entire mediolateral extent of the cerebellum in *Prdm13*^*+/+*^ mice (Supplemental Figure 7Q-T). In contrast, no PAX2+ cells were present in the dorsal region of the VZ in the cerebellar vermis and hemispheres of *Prdm13*^*-/-*^ mice, with only a few cells present in the most ventral aspect of the cerebellum (Supplemental Figure 7U-X). Quantification of PAX2+ cells at subsequent stages of development indicated that the number of PAX2+ progenitors was significantly reduced in *Prdm13*^*-/-*^ cerebella (Figure 6A). A previous study has shown that *Prdm13* deficiency in the spinal cord is associated with a reduction in GABAergic PAX2+ progenitors and an accompanying increase in glutamatergic progenitors (37). To determine if PRDM13 has a similar role in suppressing glutamatergic TLX3+ cell fate in the developing cerebellum, TLX3+ progenitors were visualized by immunostaining and quantified. At E12.5, TLX3+ cells are restricted to the most ventral aspect of the cerebellum, in all regions apart from the most lateral cerebellum where they are completely absent (Supplemental Figure 7I-L). Interestingly, in *Prdm13*^*-/-*^ mutants, TLX3+ cells expanded dorsally to occupy the majority of the dorsal-ventral extent of the cerebellum almost extending to the level of the rhombic lip. These findings are identified across the entire mediolateral cerebellum in *Prdm13*^*-/-*^ mutants (arrowheads, Supplemental Figure 7M-P). At E14.5, few TLX3+ neurons are normally present outside of the EGL in wild-type mice (Figure 6B). However, TLX3+ cells are clearly present in the *Prdm13*-deficient cerebellum just outside the ventricular zone where PAX2+ cells are normally found (Figure 6C). Quantification confirmed the reduction in PAX2+ progenitors and a concomitant increase in TLX3+ cells (Figure 6D). We subsequently asked if the mis-specified TLX3+ or PAX2+ cells accounted for the increase in cell death in the non-EGL mutant cerebella at birth. The proportion of PAX2+ and TLX3+ apoptotic cells were calculated after combining immunolabelling with a TUNEL assay. This analysis failed to identify any PAX2+ or TLX3+ cells undergoing apoptosis in either genotype suggesting that *Prdm13* is not required for the survival of these cell types in the cerebellum at birth. Together these data identified a central role for PRDM13 in the specification of GABAergic PAX2+ cells by suppressing glutamatergic cell fate.

**Figure 6.**
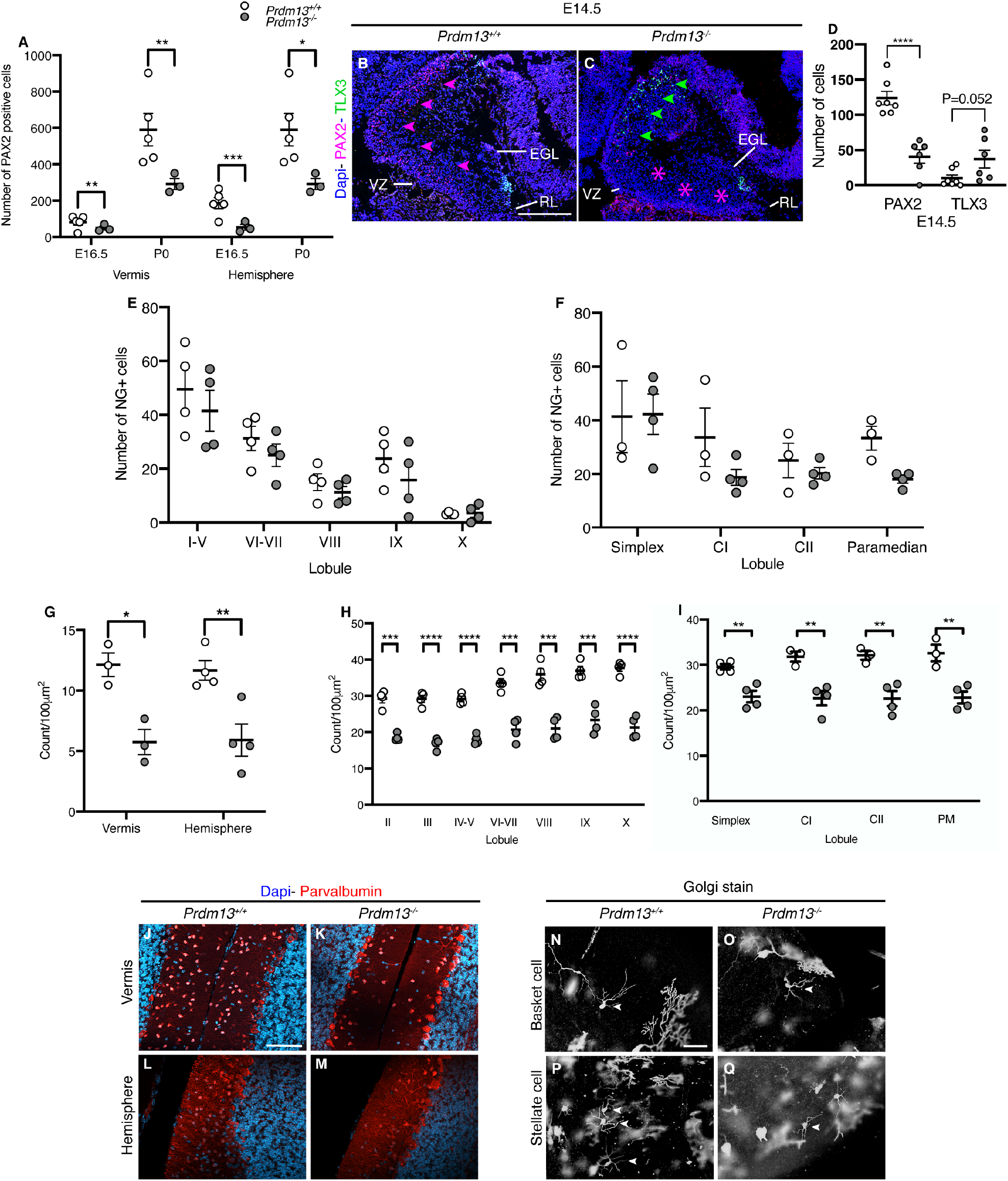
PRDM13 is a critical regulator of GABAergic cell fate in the cerebellum. **(A)** PAX2 cell counts in *Prdm13*^*+/+*^ and *Prdm13*^*-/-*^ cerebella at indicated stages. Note the reduction in PAX2 positive cells in the cerebellar vermis and hemisphere of *Prdm13*^*-/-*^ mice at E16.5 and P0 (n=4 per genotype). **(B**,**C)** Immunohistochemistry of sagittal sections through the cerebellum at E14.5 of *Prdm13*^*+/+*^ and *Prdm13*^*-/-*^ mice using antibodies to PAX2 and TLX3 to label GABAergic interneurons and glutamatergic progenitors, respectively. Note the marked reduction in PAX2 positive cells in *Prdm13*^*-/-*^ mice, indicated by pink asterisks (C) and the concomitant increase in TLX3-positive cells (C) indicated by green arrows. **(D)** Quantification of PAX2 and TLX3 positive cells is shown at E14.5. Note the decrease in PAX2+ cells is accompanied by an increase in TLX3 positive neurons. **(E**,**F)** Neurogranin+ Golgi cell counts in *Prdm13*^*+/+*^ and *Prdm13*^*-/-*^ vermis (E) and hemispheres (F) at P21. Note the similar number of Golgi cells between genotypes. **(G)** Quantification of parvalbumin+ molecular layer interneurons (MLIs) in the cerebellum at P21. Note the reduction in MLIs in the cerebellar vermis and hemispheres of *Prdm13*^*-/-*^ mice. **(H**,**I)** Lobule specific MLI cell counts in the cerebellar vermis and hemispheres at P21. Note the significant reduction in MLIs across all cerebellar vermis and hemisphere lobules of *Prdm13*^*-/-*^ cerebella. **(J-M)** Immunohistochemistry of sagittal sections through the cerebellum at P21 of *Prdm13*^*+/+*^ and *Prdm13*^*-/-*^ mice using antibodies to parvalbumin to label MLIs. Note the striking reduction in parvalbumin+ cells in the cerebellar vermis (K) and hemispheres (M) of *Prdm13*^*-/-*^ mice. **(N-Q)** Golgi-Cox stained sagittal sections of adult *Prdm13*^*+/+*^ and *Prdm13*^*-/-*^ cerebella. Note that the morphology of the Basket (N,O) and Stellate cells (P,Q) is consistent between *Prdm13*^*+/+*^ and *Prdm13*^*-/-*^ mice. *P<0.05, **P<0.01, ***P<0.001, Student’s t test. VZ = ventricular zone, EGL = external granular layer, RL = rhombic lip, NG = neurogranin, CI = Crus I, CII = CrusII. Scale bar = 300μm (B), 100μm (J, N).

The cerebellar cortex is comprised of three distinct layers where highly stereotyped connections between neurons form a repeating and relatively simple cerebellar circuit. PAX2+ precursors form a diverse population of GABAergic interneurons, which integrate into and regulate the output of all levels of the cerebellar circuit through local inhibition. To determine the impact of PAX2+ mis-specification in *Prdm13*^*-/-*^ mutants, we analysed the two major GABAergic interneuron populations of the cerebellar cortex, Neurogranin^+^ Golgi cells and Parvalbumin^+^ molecular layer interneurons (MLIs). Golgi cells were distributed appropriately throughout the granular cell layer with normal cell number in all lobules of the cerebellar vermis and hemispheres of *Prdm13*-deficient cerebella (Figure 6E,F). In contrast, MLIs, confined to the molecular layer of the cerebellar cortex, were significantly reduced in *Prdm13*^*-/-*^ mice (Figure 6G,J-M). A finer analysis revealed that this reduction was uniform across all cerebellar lobules of the vermis and hemispheres (Figure 6H,I). To determine if this alteration in MLI number was associated with any changes in morphology, we performed a Golgi-Cox stain in adult mice. There are two recognized subtypes of MLI, Basket and Stellate cells, characterised according to location in the molecular layer and morphology. Small somata of Basket and Stellate interneurons were respectively identified in the inner and outer portions of the molecular layer (Figure 6N-Q). No morphological change in these neurons was observed in *Prdm13*^*-/-*^ mutants. Specifically, Basket cells had long, straight dendrites, that extended towards the pial surface with a single, main axon that ran parallel to the PC layer (Figure 6O). Stellate cells were identifiable by their characteristic star-like appearance created by radiation of dendrites from the cell body (Figure 6Q).

Together these findings suggest that mis-specification of PAX2+ cells results in fewer MLIs in the adult cerebellum. As MLIs are postulated to influence cerebellar-regulated behaviours by regulating PC activity (52, 55-57) deficits in MLIs may underlie some of the neurological symptoms observed in individuals with homozygous *PRDM13* mutations.

## Discussion

Here we describe a novel, recessive syndrome most likely caused by a hypomorphic mutation of *PRDM13*, present at low levels in the same ethnic background and inherited from unaffected, heterozygous parents. Patients homozygous for this mutation exhibited a constellation of phenotypes that included developmental delay, CHH, cerebellar hypoplasia, scoliosis, and intellectual disability. To understand how PRDM13 deficiency might lead to CHH and cerebellar hypoplasia, we studied hypothalamic and cerebellar development in a homozygous *Prdm13*-deficient mouse model. This analysis identified key roles for PRDM13 in the development of Kiss1 neurons in the hypothalamus and molecular layer interneurons in the cerebellum. Intriguingly, both these neuronal populations also require the transcription factor PTF1A for normal development, and since PTF1A has been shown to control *Prdm13* expression in the developing cerebellum and spinal cord, our findings suggest that the CHH and cerebellar phenotypes are functionally linked by central roles for PTF1A and PRDM13 in neuronal cell fate specification in these distinct tissues.

### Recessive PRDM13 mutation is associated with a unique combination of phenotypes

The combination of developmental delay, CHH, cerebellar hypoplasia, scoliosis, and intellectual disability associated with *PRDM13* mutation differs significantly from other conditions with CHH and cerebellar hypoplasia, such as Gordon Holmes syndrome (GHS). The latter is an autosomal recessive adult-onset neurodegenerative disorder characterized by progressive cognitive decline, dementia, and variable movement disorders, such as ataxia and chorea. Mutations in genes encoding for ubiquitin ligase and deubiquitinase such as *RNF216, OTUD4* and *STUB1* have been identified in GHS patients (10, 58-61), linking disordered ubiquitination to neurodegeneration and reproductive dysfunction. *PNPLA6* mutations are associated with Boucher-Neuhauser, Oliver McFarlane and Lawrence-Moon syndromes, in which CHH and progressive cerebellar ataxia may be prominent features (62-64). In contrast to the late-onset neurodegenerative etiology of cerebellar atrophy in these conditions, we show here that *PRDM13* mutations disrupt cerebellar development.

### The PTF1A-PRDM13 axis in Kiss1 neuron development

Previous studies showed that PRDM13 functions downstream of PTF1A, which plays distinct roles in the regulation of both the cerebellum and the hypothalamus (20, 21, 40). Our observations support the existence of overlapping phenotypes between *Prdm13*- and *Ptf1a*-deficient mice in the hypothalamus and cerebellum, implying that a transcriptional dysregulation in specific neuronal progenitors is the most likely pathogenic mechanism underlying the phenotypes associated with *PRDM13* mutation. Although *PTF1A* mutations have been linked to cerebellar aplasia and hypoplasia, no link to CHH in humans has, to our knowledge, been reported to date. Defective Kiss1 neuron development has been described in mice with conditional forebrain-specific *Ptf1a* deletion using the Nkx2.1-Cre driver (40). Interestingly, in this study, Kiss1 neurons were found not to be in the *Ptf1a* lineage. Thus, PTF1A induction of *Prdm13* expression in the same lineage cells could instruct neighbouring cells to differentiate into Kiss1 neurons via transmembrane and/or secreted proteins (40, 65). Members of the semaphorin family were among the genes affected by PTF1A loss in the forebrain (40). Given their key roles in the control of the reproductive axis (66), it will be interesting to explore their possible involvement in the differentiation program of Kiss1 neurons, thus shedding light into the cellular mechanisms through which the PTF1A-PRDM13 axis regulates reproduction.

While patients carrying *PRDM13* mutations present with CHH, we did not find typical phenotypes of hypogonadism in *Prdm13*-deficient mice, except for a delayed pubertal onset in female mice. Importantly, although we were not able to detect Kiss1 neurons in E14.5 *Prdm13*-deficient mice, some Kiss1 neurons were present in the ARC nucleus of adult *Prdm13*-deficient mice. Previous studies have shown that a residual 5% function of Kiss1 neurons is sufficient to guarantee male mouse fertility (67), and that mice lacking *Kiss1* display a variable reproductive phenotype (50). Thus, our data suggest that sufficient numbers of Kiss1-expressing cells differentiate in *Prdm13*^*-/-*^ mutants to support normal sexual development, whereas the reduced number may have an impact on pubertal onset, causing a delay.

Our data highlight that PRDM13 selectively controls a subset of hypothalamic neurons, underlining a cell-specific role of PRDM13 in the differentiation program of the hypothalamus. In contrast, despite an overall downregulation of *Gad1* in the hypothalamus of *Prdm13*-deficient mice, the expression of typical hypothalamic GABAergic neuronal markers was unaltered Yet, given that GABAergic neurotransmission at the hypothalamic level is on its own fundamental in sustaining fertility and pubertal onset (45), reduced levels of GABA might also contribute to CHH observed in the patients.

Interestingly and in contrast to *Ptf1a*-deficient mice, our data also indicate that *Prdm13* deficiency specifically affects Kiss1 neurons in the Arc nucleus, but not in the AVPV, suggesting a highly restrictive role for PRDM13 in the specification of Arc Kiss1 neurons and the possible involvement of additional transcription factors in the specification of AVPV Kiss1 neurons. These observations may also explain the inability to detect a significant reduction in *Kiss1* expression in the whole hypothalamic region of female *Prdm13*-deficient mice (Figure 4D), as unaffected AVPV Kiss1 neurons are more dense in females than in males (48).

To our knowledge, this is the first evidence linking disrupted PRDM13-mediated regulation of Kiss1 neurons to CHH in humans. It would be interesting to assess the effects of Kisspeptin administration on gonadotropin secretion in our mouse model, leading to a potential treatment for patients that carry a *PRDM13* mutation.

### PRDM13 as a key GABAergic fate regulator in the cerebellum

The molecular mechanisms that control the specification, maintenance and differentiation of GABAergic progenitors from a common PTF1A+ progenitor pool in the cerebellar ventricular zone remain ill-defined. We have identified a clear role for PRDM13 in controlling GABAergic fate in the developing cerebellum. As *Prdm13* expression requires PTF1A in the cerebellum (37), we conclude that PRDM13 functions as a critical effector protein downstream of PTF1A in GABAergic cell fate regulation. Specifically, we demonstrate a differential requirement for PRDM13 in GABAergic neuronal development, where PRDM13 is necessary to generate a subset of PAX2+ GABAergic interneurons, but appears largely dispensable for PC development. Previous analyses of *Prdm13* in the dorsal neural tube suggested that the *Prdm13* phenotype is less severe than that observed in *Ptf1a* mutants (37). Indeed, there is an absolute and global requirement for PTF1A in GABAergic neuronal specification, and PTF1A deficiency leads to cerebellar agenesis (20). It will be of interest to assess cerebellar and hypothalamic phenotypes in *Prdm13* conditional mutants to determine whether the milder phenotype in our *Prdm13* mutants is because PTF1A regulates other genetic pathways in addition to *Prdm13*, or whether the milder phenotype is merely a consequence of residual PRDM13 function in our *Prdm13* mutants. The same holds true for the sexual differentiation phenotypes.

More broadly, these data implicate PRDM13 in the regulation of excitatory:inhibitory balance in the cerebellum. GABAergic PAX2+ interneuron progenitors are lost in *Prdm13*-deficient mice, and progenitors expressing the glutamatergic marker TLX3 appear in their place, thus, there is a change in cell fate in the absence of *Prdm13*, similar to findings in *Ptf1a* mutants (25). This is particularly interesting when considering that all patients had moderate to severe intellectual disability. Cerebellar dysfunction and altered cerebello-cortical circuitry is thought to contribute substantially to complex neuropsychiatric diseases and has been linked to autism (68), intellectual disability and schizophrenia (69, 70). One pathophysiological theory linking these genetically heterogenous and diverse neuropsychiatric disorders is one of altered excitatory:inhibitory balance (71), a theory that has been gaining significant traction. It is interesting, therefore, that Patient 3 also developed epileptic seizures, adding further weight to the possibility of altered excitatory:inhibitory balance in patients.

To conclude, we have identified a novel, recessive syndrome associated with a mutation in the *PRDM13* gene. Our analysis of mice homozygous for a hypomorphic *Prdm13* allele suggests that the phenotypic association of CHH with cerebellar hypoplasia occurs as a result of the conserved regulation of GABAergic and Kiss1 neuronal differentition in the cerebellum and hypothalamus respectively, by PRDM13 downstream of the transcriptional regulator PTF1A. Patients with recessive, hypomorphic *PRDM13* and/or *PTF1A* mutations are likely to be extremely rare; however, interestingly, the three patients, and the unrelated heterozygous carrier with the 13bp deletion in *PRDM13*, shared an identical 0.2Mb region of homozygosity that encompassed the *PRDM13* mutation, which extended for 1.6Mb in total in the three patients (Supplemental Table 3). This suggests that the *PRDM13* mutation is present on a common haplotype within the same population and may in turn lead to identification of further patients with this phenotype. The existence of patients with *PRDM13* mutations and the associated phenotypes reported here in *Prdm13*-deficient mice represent an important step towards dissecting the molecular mechanisms controlling GABAergic fate determination in the developing nervous system. These mechanisms are likely to contribute to excitatory:inhibitory imbalance in the brain, which may underlie a range of different neurodevelopmental phenotypes.

## Methods

### Whole exome and Sanger sequencing

Whole exome capture was performed on all family members using Agilent SureSelect version 4 (Santa Clara, CA), according to manufacturer’s protocol. Enriched libraries were sequenced on the Illumina (San Diego, CA) HiSeq2500. Sequencing reads passing quality filters were aligned to the reference genome build GRCh37/hg19 using Burrows-Wheeler Aligner (BWA) algorithm and for variant calling we applied GATK (72) base quality score recalibration, indel realignment, duplicate removal, and performed SNP and INDEL discovery and genotyping using standard hard filtering parameters or variant quality score (73). The variant annotation and interpretation analyses were generated using Ingenuity® Variant Analysis™ (version 4.0.20151113) software from Qiagen (Redwood City, CA). Disease-causing variants were validated by Sanger sequencing.

### Exome sequencing filtering strategy

Exonic and cryptic splice site variants (+/-5), which were homozygous in Patients 1 and 2 and heterozygous in the parents respectively, had a call quality ≥20, read depth ≥10 and had a frequency of <0.1% in public exome databases (1000 genomes, NHLBI ESP exomes, ExAC) or were known pathogenic variants listed in HGMD (Human Genome Mutation Database). Initially, Kallmann syndrome, hypogonadotropic hypogonadism (HH), cerebellar hypoplasia, developmental delay, or diseases consistent with these phenotypes were set as the biological terms in our filtering criteria.

### Microarray

Four samples; the three affected patients and an unrelated heterozygous carrier of the 13bp deletion in PRDM13 were genotyped using an Illumina Infinium OmniExpress-48 microarray according to the manufacturers’ instructions and processed using Illumina software.

### Ethics

The appropriate ethical approval for the genetics and human embryonic tissue expression studies has been obtained prior to this project taking place. The human embryonic and fetal material was provided by the Joint MRC/Wellcome Trust (grant# MR/R006237/1) Human Developmental Biology Resource (HDBR) (http://hdbr.org). Ethical committee approval for study of patient DNA samples was obtained from the Institute of Child Health/Great Ormond Street Hospital for Children Joint Research Ethics Committee. Informed consent was obtained from the parents of the patients prior to collection of samples, genomic analysis and publication.

### Mice

*Prdm13* mutant mice lacking exon 2 and 3, which encode much of the PR domain, have been described and maintained on a C57BL/6 background (35). Mice were genotyped by PCR using DNA obtained from the ear as described in the original publication (See Supplemental data for primer sequences). All mice were maintained and bred in the Biological Services Unit at Guy’s Campus, with all experiments approved by the local ethical review board and the UK Home Office (Project licence P8DC5B496).

### Histology

Samples were dissected in PBS and fixed in 4% paraformaldehyde overnight at 4°C and either cryopreserved in 30% sucrose for OCT embedding or, after dehydration, infiltrated and embedded in paraffin wax. For samples immunostained for TLX3, dissected samples were cryoprotected in 7.5% gelatin and 15% sucrose. Serial sagittal sections were cut at 10 μm before drying at 42°C overnight. Sagittal or coronal cryosections were cut at 10-20 μm using a Cryostat. *0*.*1% Cresyl violet acetate (Nissl) staining*. Sections were deparaffinised in xylene, rehydrated through graded ethanol solutions and stained with 0.1% Cresyl Violet Acetate for 10 minutes. Differentiation of the stain was achieved using Glacial Acetic Acid. Stained sections were dehydrated, placed in xylene and mounted.

### Haematoxylin and Eosin staining

Sections were deparaffinised in xylene and rehydrated in graded ethanols. Sections were stained with Erhlich’s Haematoxylin for 10 minutes, washed to remove excess stain and immersed in acid alcohol (0.5%HCl, 70% ethanol) for 15 seconds. Sections were then stained with 0.5% aqueous Eosin for 2 minutes. Stained sections were dehydrated, placed in xylene and mounted.

### Immunohistochemistry

Immunohistochemistry on paraffin or cryosections was performed using standard methods (74, 75). Antibodies were: guinea pig anti-PRDM13 (1:1000; described in (35)), rabbit anti-Purkinje cell protein 2 (anti-PCP2) (1:200; gift from Brad Denker, Harvard University, Boston, Massachusetts, USA), mouse anti-LHX1,5 (1:100; Hybridoma bank 4F2), rabbit anti-PAX2 (1:200; ThermoFisher 71-600); Guinea pig anti-TLX3 (1:10000; gift from Thomas Muller, MDC Molecular Medicine), anti-cleaved caspase 3 (Asp 175) (1:150; Cell Signaling Technology 9661), anti-phosphohistone H3B (Ser 10) (1:200; Abcam Ab14955), rabbit anti-GnRH (1:400; ImmunoStar 20075), rabbit anti-SOX9 (gift from Prof. Wegner), rabbit anti-Neurogranin (1:100; Merck Millipore AB5620), rabbit anti-Parvalbumin (1:200; Abcam AB11427). Sections were incubated with Alexa Fluor labelled secondary antibodies (1:200; Life Technologies) and counterstained with DAPI to allow detection of PRDM13, PCP2, PAX2, TLX3, Neurogranin, Parvalbumin and PH3B. Fluorescent images were acquired from Citifluor (Citifluor Ltd.) mounted slides using a Nikon Eclipse 80i microscope with a Nikon Y-QT Hamamatsu C4742-95 camera. For PAX2, TLX3, cleaved-caspase 3, GnRH and SOX9 detection, sections were incubated with species-specific biotinylated secondary antibody (1:200; Dako) for 1-2 hours. VECTASTAIN Avidin/Biotin Complex (ABC) kit (1:200; Vector Laboratories Ltd.) was used to amplify the signal prior to visualisation. Sections were incubated in 3-3’-diaminobenzidine substrate (0.03%; Sigma-Aldrich), where needed dehydrated, mounted and visualised with a Nikon Eclipse 80i microscope. For detection of TLX3, cryosections were processed for antigen retrieval using 0.2% Triton X-100/PBS at 40°C for 20 minutes.

### In situ hybridisation

*In situ* hybridization on mouse sections was performed using previously described methods (76). DNA templates were generated from mouse genomic DNA by PCR amplification. The PCR primers used were: *Prdm13* (exon4) forward 5’-GCCACTTGTGCCTCTACTGT-3’, reverse 5’-CCTCCACAGACAAGAGCGTT-3’. The T7 promoter was added at 5’ of the reverse primer. A DIG RNA labeling kit with T7 RNA polymerase (Roche) was utilized to produce a *Prdm13* anti-sense probe by *in vitro* transcription. *In situ* hybridisation on human tissue sections at CS23 of human embryonic development was carried out as previously described (77). The sections were prepared by the Human Developmental Biology Resource (HDBR) and a purified vector containing a conserved portion of human wild-type *PRDM13* cDNA (Source Bioscience) was used to make the digoxigenin (DIG)-labelled (Promega) RNA probes.

### RT-PCR and quantitative RT-PCR

Total RNA was extracted from samples (three of each genotype) using TRIzol (Life Technologies) and Direct-zol RNA miniprep kit (Zymo Research). cDNA was synthesized from 200ng of total RNA using nanoScript2 Reverse Transcription kit (Primer design Ltd) and random hexamer primers. Conventional PCR and analysis by gel electrophoresis on samples was performed as previously described (78). Quantitative-PCR was performed on a ROCHE LightCycler 480 qPCR machine using the precision qPCR master mix with SYBR Green (Primerdesign Ltd.). Triplicate samples were run in all reactions; first-strand DNA synthesis reactions without reverse transcriptase were used as controls. The ΔCq value and the ΔΔCq were calculated relative to control samples using quantification cycle (Cq) threshold values that were normalised to the housekeeping gene, *Gapdh*.

Primer sequences used are in Supplemental data.

### Statistics

Statistical tests employed are outlined in figure legends and were conducted when the experiment had been performed a minimum of 3 times on a minimum of 3 individual samples. Data are presented as mean ± SEM and results considered significant with a *P* value less than 0.05.

### Study approval

Animal housing and experimental procedures complied with the local ethical review panel of King’s College London, and the UK Home Office Animals Scientific Procedures Act 1986. The work was performed under project licences (PPL70/6694, PPL70/7184 and P8DC5B496).

### PAX2, TLX3, cleaved caspase 3, phosphohistone H3, Parvalbumin and Neurogranin counts

Counts were performed on IHC labelled cells on serial sagittal sections. The total number of labelled cells were counted using ImageJ (NIH) on images obtained using a Zeiss ApoTome microscope.

### GnRH neuron counts

Counts were performed as previously described (75, 79).

### Area analysis

The surface area of Cresyl-violet (0.1%) serial sagittal sections was measured using ImageJ (NIH) and used to estimate the cerebellar area.

### Molecular layer thickness

Molecular layer thickness measurements were performed in the lobule III/IV region using PCP2 labelled sections as previously described (52).

### Golgi- Cox staining

Golgi- Cox staining was performed with the FD Rapid Golgi Stain Kit (FD Neurotechnologies PK401) on adult brain samples, according to the manufacturer’s instructions. Following staining, sections were dehydrated, cleared with xylene and mounted. Images were obtained using a Leica DMI6000 microscope.

### Behaviour – motor coordination and learning

Motor coordination and learning were assessed from 42 days of age on an accelerating rotarod (Panlab Harvard Apparatus) as described previously (80).

### Puberty assessment

*Prdm13*^*+/+*^ and *Prdm13*^*-/-*^ female mice were checked daily for vaginal opening as previously described (81).

## Data Availability

Requests for raw data and materials should be made to the corresponding authors. No large data sets form part of the manuscript.

## Author contributions

MD, HJW, MAB and AC conceived the project and coordinated and supervised experimental work. LCG performed the human expression studies, generated the table of clinical patient data.

JT, NF, MJC, MD recruited the patients to the study and performed phenotypic characterisation of the patients.

HJW, PLQS, GOSgene and LO performed the next generation sequencing analysis, variant interpretation and Sanger sequencing of all participants.

TC and TF provided *Prdm13*-deficient and control tissues and animals and PRDM13-specific antiserum. DEW, KLR and MAB established and maintained the *Prdm13* mouse colony at KCL. DEW, DF, SR and KLR performed the analysis of cerebellar development.

RO, AL, LBD, AJJP performed the analysis related to the GnRH- and Kiss1-neuronal systems, included the histological analyses of gonads. RO and AC prepared figures 1-4 and Supplemantal Figure 2.

DEW, RO, LCG, ICAFR, AC, MAB and MD wrote the manuscript with contributions from all authors.

## Acknowledgments

We thank Drs. Thomas Muller and Brad Denker for the TLX3 and PCP2 antiserum, respectively, and Dr. Laura Croci for technical assistance with Kiss1 *in situ* hybridization. We thank Dr. Minaxi Dattani for her help with the preparation of Figure 1. We are also grateful to Professor Christiana Ruhrberg for access to equipment and reagents during the initial phase of the project.

RO was supported by a short-term fellowship from EMBO (number 7950) to visit the Basson lab. DEW was supported by an Integrated Training Fellowship from the Wellcome Trust (WT096385MA), short-term fellowship support from the Royal Veterinary College and King’s College London and a Starter Grant for Clinical Lecturers (SGL023_111). MTD receives funding from the Great Ormond Street Hospital (GOSH) Children’s Charity and the Medical Research Foundation, UK (grant# 535963). Research at GOSH benefits from funding received from the NIHR Biomedical Research Centre. AC was funded by the Italian Ministry of Health (GR-2016-02362389). TF was funded by Grant-in-Aid for Scientific Research (18H02593) from the Japan Society for the Promotion of Science and The Takeda Science Foundation.

## Figures

**Supplemental Figure 1.**
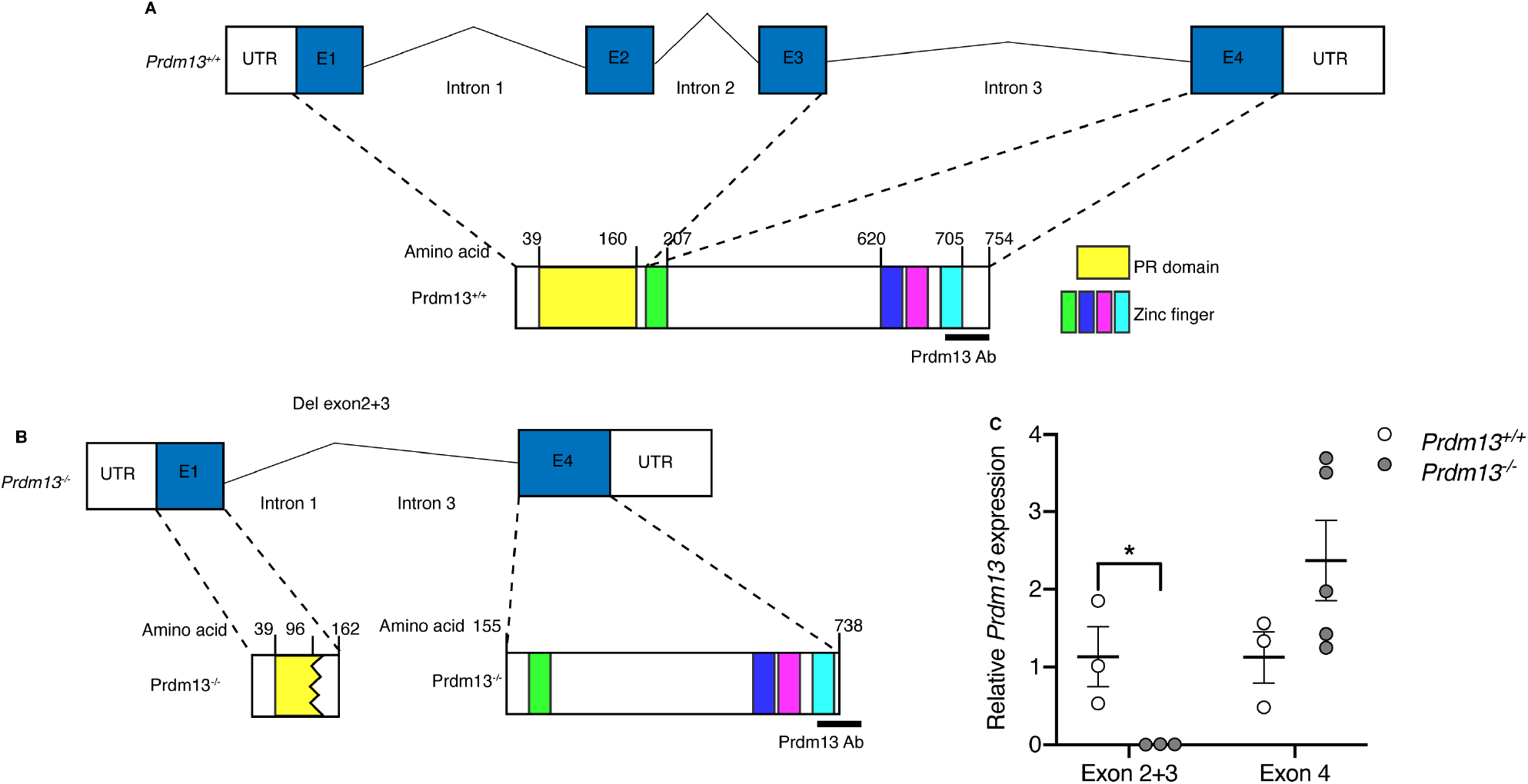
*Prdm13*^*Δex2,3*^*/* ^*Δex2,3*^ mutant allele. **(A)** Schematic representation of the mouse *Prdm13* locus and protein. The untranslated regions (UTR) are labelled. Exons are indicated by blue boxes and labelled E1, E2, E3 and E4. The functional domains encoded by the exons are shown and labelled according to the key provided. Dashed lines indicate the regions of the protein encoded by exon 1-3 and exon 4. (B) Schematic representation of the mouse *Prdm13* locus and the predicted *Prdm13*^*-/-*^ protein. Predicted translated proteins are indicated and the C-terminal epitope of antiserum used to identify the wild-type protein is shown. (C) Quantification of *Prdm13* exon 2+3 and exon 4 transcript levels in E12.5 cerebella. Note the significant reduction in exon 2+3 expression in *Prdm13* mutants whilst exon 4 transcripts were increased but did not reach significance. *P<0.05, **P<0.01, Student’s *t* test. E1 = exon1, E2 = exon2, E3 = exon3, E4 = exon4, UTR = untranslated region.

**Supplemental Figure 2.**
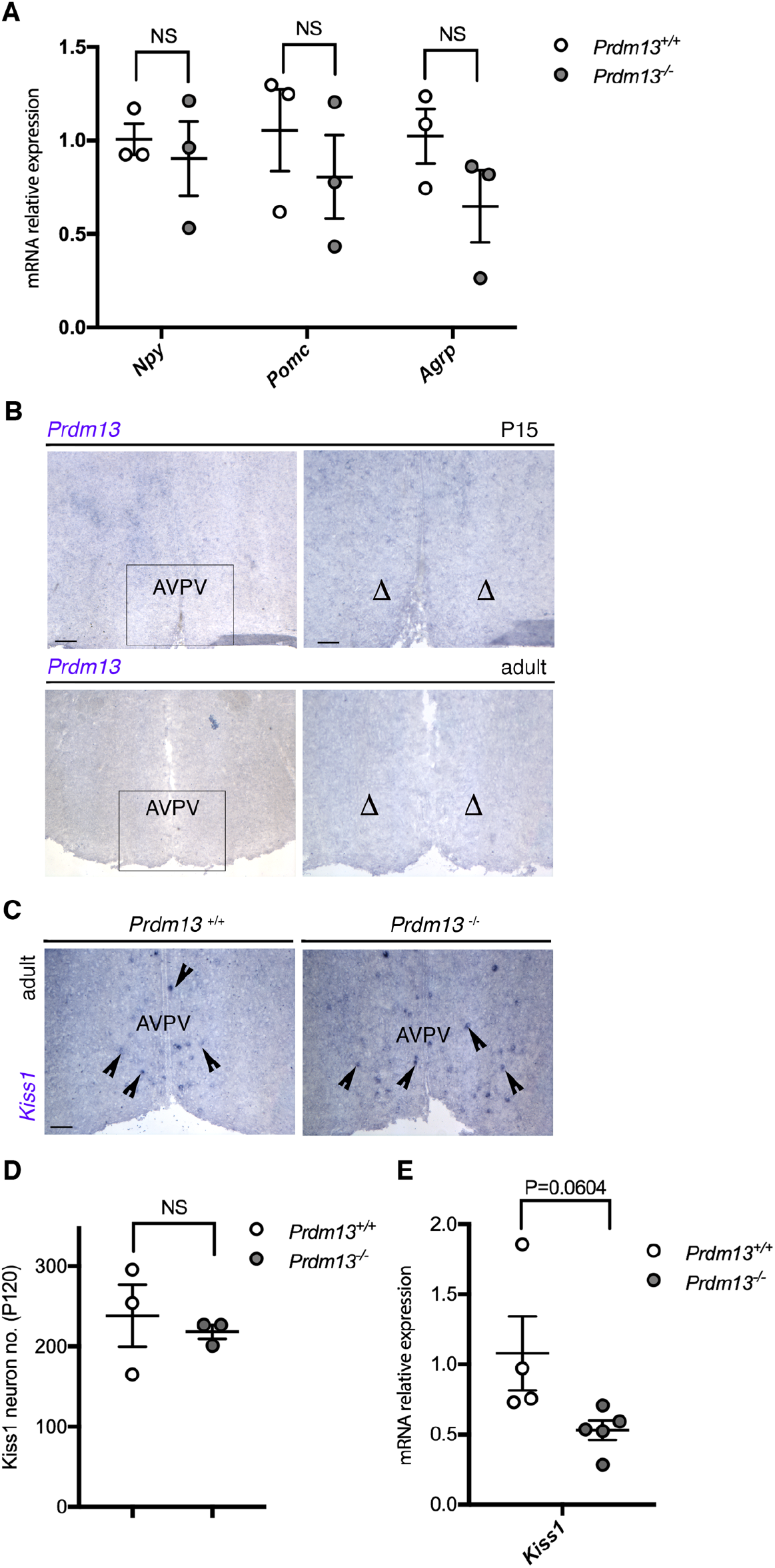
*Prdm13*-null mice display similar levels of Arc GABAergic neuronal markers and a similar number of AVPV Kiss1 neurons compared to wild-type mice. **(A)** qRT-PCR analysis for *Npy, Pomc, Agrp* transcripts in the hypothalamus of *Prdm13*^*+/+*^ and *Prdm13*^*-/-*^ male mice. ΔΔCq were calculated relative to control samples using quantification cycle (Cq) threshold values that were normalised to the housekeeping gene, *Gapdh*. **(B)** *In situ* hybridization experiments on coronal P15 and adult male brain sections to detect the expression of *Prdm13* in the AVPV nucleus of the hypothalamus. High magnification of the squared areas are shown next to each panel. Note the absence of *Prdm13* transcript expression in the AVPV nucleus at both stages (Δ). **(C)** *In situ* hybridization experiments on sections taken at the level of the AVPV nucleus from *Prdm13*^*+/+*^ and *Prdm13*^*-/-*^ adult male mice for *Kiss1* transcripts. **(D)** Quantification of Kiss1-positive neurons in the AVPV nucleus from *Prdm13*^*+/+*^ and *Prdm13*^*-/-*^ adult male mice. No differences in *Kiss1* expression were observed in the mutants compared to wild-types. Arrowheads indicate *Kiss1*-positive neurons. **(E)** qRT-PCR analysis for *Kiss1* transcript in the hypothalamus of *Prdm13*^*+/+*^ and *Prdm13*^*-/-*^ female mice. ΔΔCq were calculated relative to control samples using Cq threshold values that were normalised to the housekeeping gene, *Gapdh*. Note the non-significantly decreased *Kiss1* levels in mutant mice compared to wild-type. Scale bars = 500 μm (B low magnification), 250 μm (B-C high magnification). AVPV = Anteroventral periventricular nucleus.

**Supplemental figure 3.**
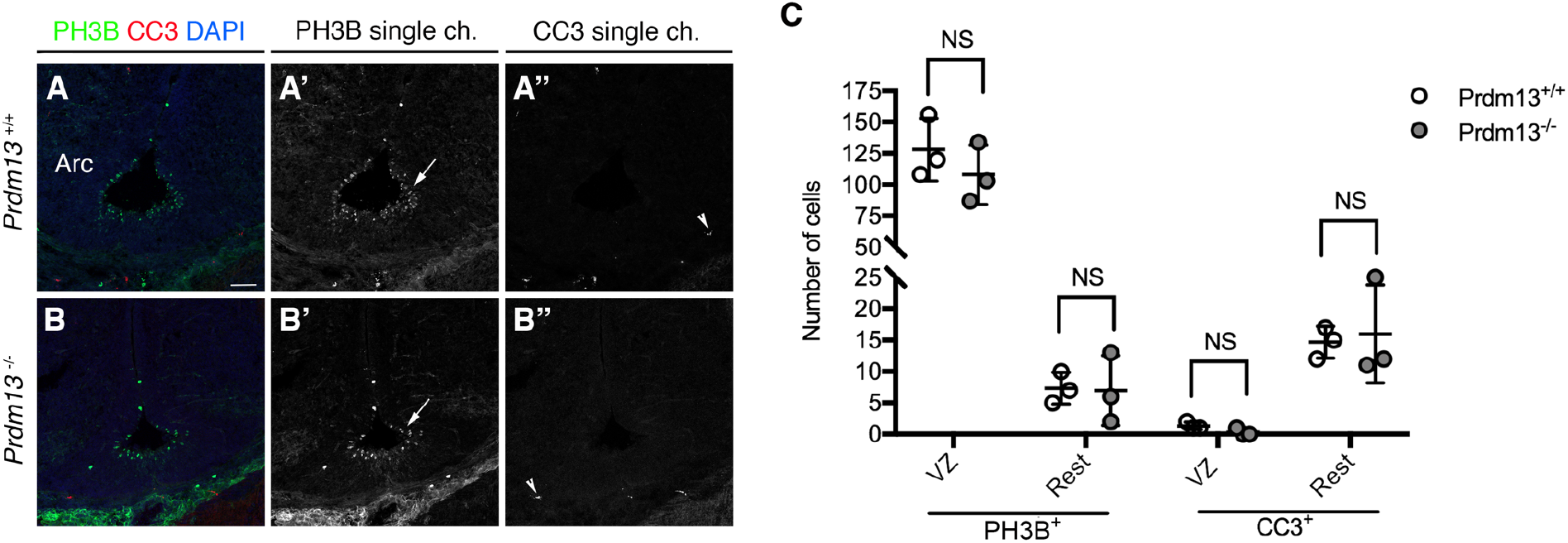
Loss of Kiss1 neurons in the developing hypothalamus of *Prdm13*^*-/-*^ mice is not associated with altered cell proliferation or cell death. **(A-B)** PH3B (green) and CC3 (red) co-immunostained representative sections from E14.5 embryos of indicated genotypes at the level of the developing Arc nucleus of the hypothalamus. Single channels are shown beside each image. Arrows indicate examples of PH3B^+^ cells; arrowheads indicate CC3^+^ cells. **(C)** Quantification of proliferating PH3B^+^ and apoptotic CC3^+^ cells in the VZ and in the rest of the developing hypothalamus. No significant difference in proliferation was noted between wild-types and mutants. Scale bar = 250μm. Arc = arcuate nucleus, VZ = ventricular zone.

**Supplemental Figure 4.**
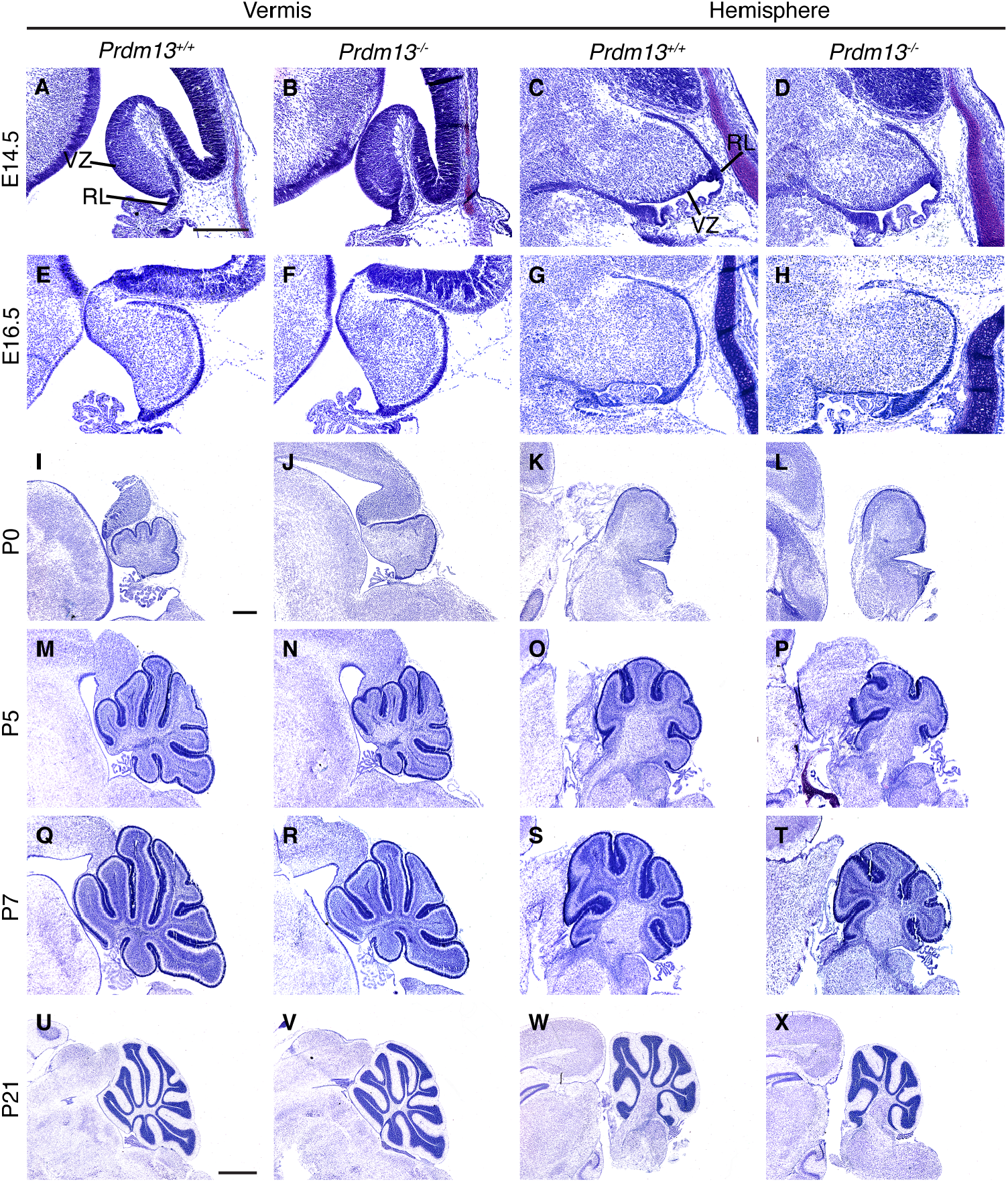
Time course of *Prdm13*^*-/-*^ cerebellar development demonstrating cerebellar hypoplasia at early postnatal stages. Cresyl violet-stained sagittal sections through the cerebellar vermis and hemisphere of *Prdm13*^*+/+*^ and *Prdm13*^*-/-*^ mice at indicated stages (A-X). Note subtle abnormalities in cerebellar vermis foliation at P0 and overt cerebellar hypoplasia in the vermis by P5 and the hemispheres by P7. VZ = ventricular zone, RL = rhombic lip. Scale bar = 300μm (A,I), 1mm (U).

**Supplemental Figure 5.**
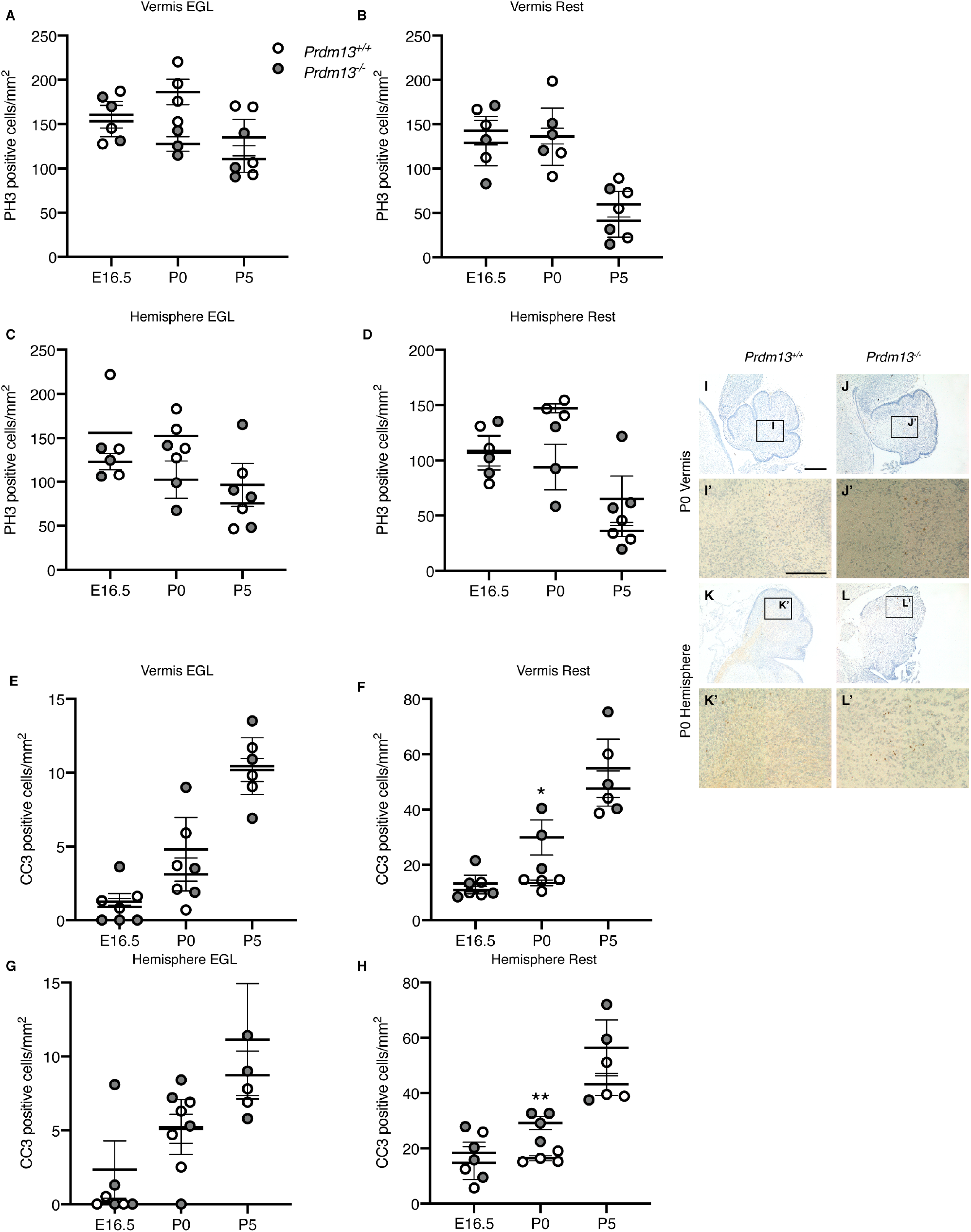
Early postnatal cerebellar hypoplasia is associated with increased apoptosis of non-EGL progenitors. **(A-D)** Quantification of phosphohistone 3B (PH3B) positive cells, in the EGL and rest of the cerebellar vermis and hemispheres as determined per mm^2^ at stages indicated (n=3 per genotype). No significant difference in proliferation was noted in the vermis or hemispheres at any stage. (E-H) Quantification of cleaved caspase 3 (CC3) positive cells, in the EGL and rest of the cerebellar vermis and hemispheres per mm^2^ at stages indicated (n=3 per genotype). Note the significant increase in apoptosis at postnatal day 0 (P0) in all but the EGL of *Prdm13* deficient mice (F,H). (I-L) Examples of CC3 immunostains, counterstained with haematoxylin, to visualise apoptotic cells on sagittal sections through the cerebellar vermis (I,K) and hemispheres (J,L) at P0, anterior to the left. Magnified views of CC3+ cells in the non-EGL cerebellum at P0 indicated by black boxes in corresponding low power views (I’,J’,K’L’). Note the increase in the number of cells undergoing apoptosis in *Prdm13* deficient cerebella (J’,L’) *P<0.05, **P<0.01, Student’s *t* test, Scale bar = 300μm (I, I’).

**Supplemental Figure 6.**
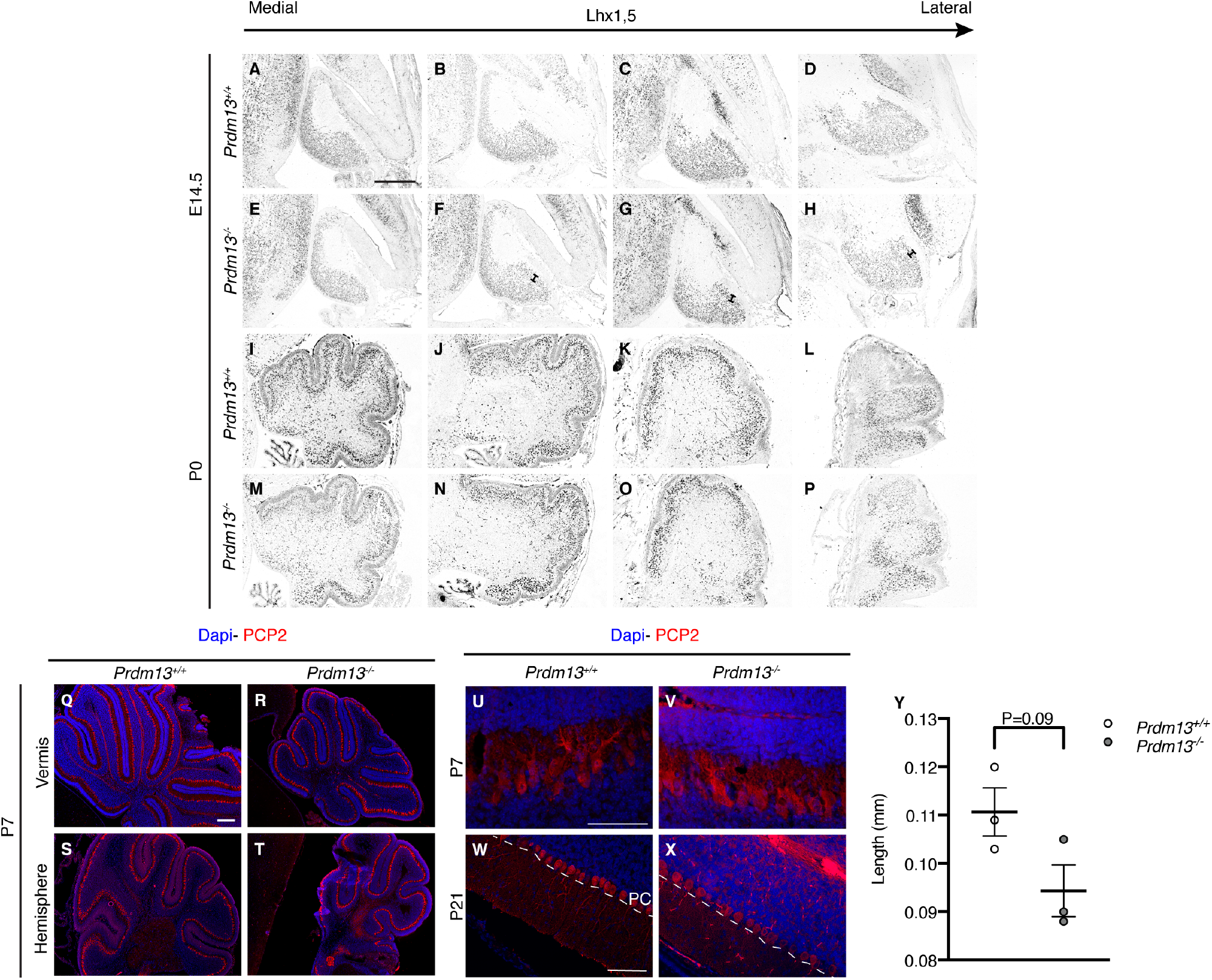
*Prdm13* is not required for PC development. LHX1,5 immunohistochemistry on sagittal sections through the cerebellar vermis and hemispheres at E14.5 (A-H) and P0 (I-P) to label PC progenitors. (Q-T) PCP2 immunochemistry on sagittal sections through the cerebellar vermis and hemispheres at P7 and P21. Note normal PC plate formation in *Prdm13*^*-/-*^ (F-H) cerebella at E14.5, indicated by brackets. Note normal PC distribution at P0 (M-P), normal PC monolayer formation at P7 (R,T) and dendritic arbor at P21 (U-X). (Y) Molecular layer width taken from lobule III/IV to estimate the span of the PC dendritic arbor (n=3 per genotype). Scale bar = 300μm (A, Q), 100μm (U,W). PC = Purkinje cell.

**Supplemental Figure 7.**
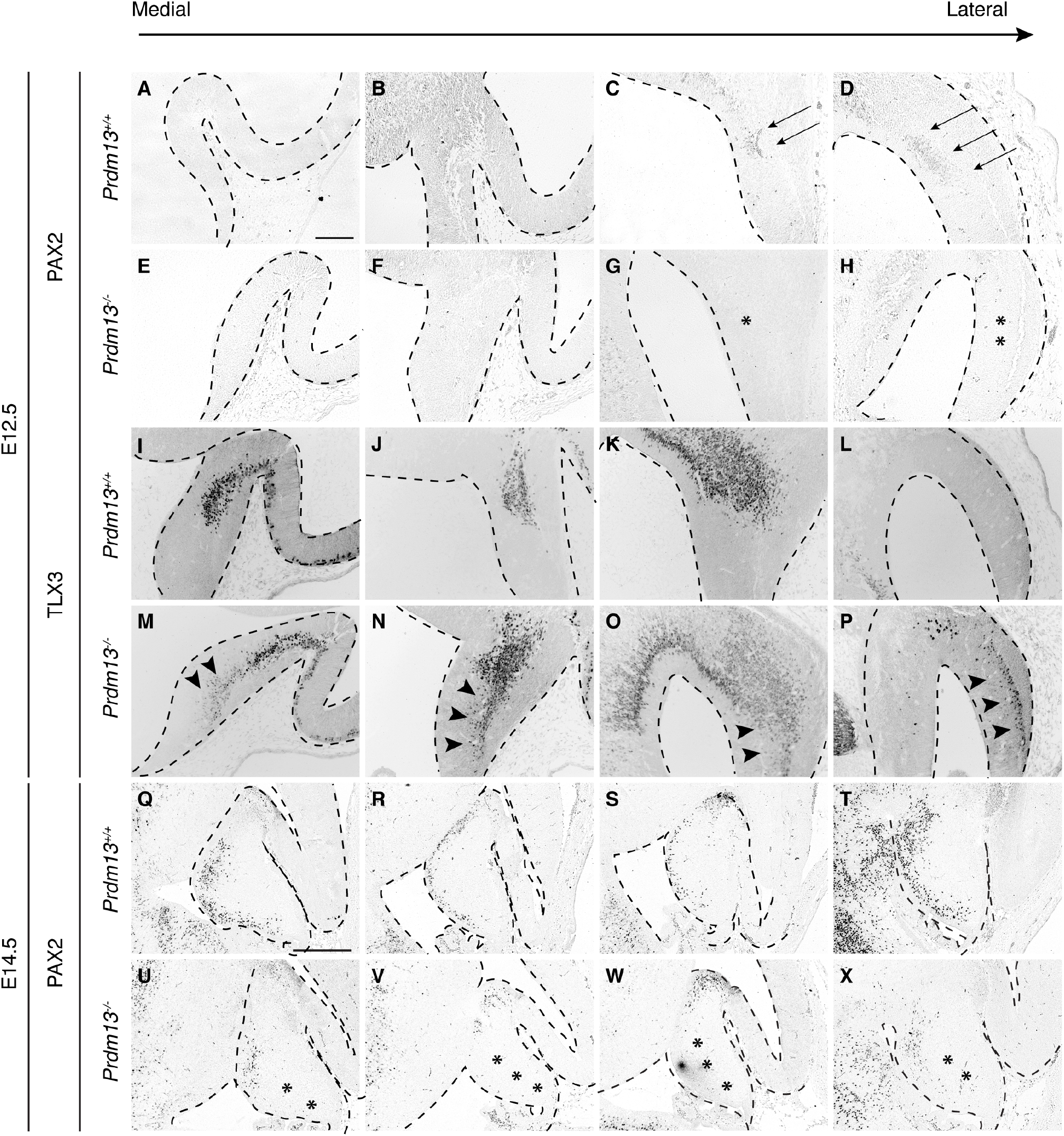
*Prdm13* is required for early GABAergic fate specification. PAX2 and TLX3 immunostains on sagittal sections through the cerebellum at stages indicated to label GABAergic interneurons and glutamatergic neurons, respectively. (A-P) Note the small cluster of PAX2+ cells in the lateral cerebellum of *Prdm13*^*+/+*^ mice at E12.5, indicated by arrows (C,D), which were absent from *Prdm13*^*-/-*^ cerebella at the same stage, indicated by asterisks (G,H). Note that at E12.5 there is expansion of the TLX3+ population laterally (P) and dorsally (M-P) where these cells occupy the length of the ventro-dorsal cerebellum in *Prdm13*^*-/-*^ mice, indicated by arrowheads (M-P). At E14.5 there is a reduction of PAX2+ neurons in *Prdm13*^*-/-*^ cerebellar vermis, indicated by asterisks (U-X). Scale bar = 300μm (A, Q).

## References

1. Boehm U, Bouloux PM, Dattani MT, de Roux N, Dode C, Dunkel L, et al. Expert consensus document: European Consensus Statement on congenital hypogonadotropic hypogonadism--pathogenesis, diagnosis and treatment. Nat Rev Endocrinol. 2015;11(9):547–64.

2. Wray S. Development of gonadotropin-releasing hormone-1 neurons. Front Neuroendocrinol. 2002;23(3):292–316.

3. Forni PE, and Wray S. GnRH, anosmia and hypogonadotropic hypogonadism--where are we? Front Neuroendocrinol. 2015;36:165–77.

4. Seminara SB, Messager S, Chatzidaki EE, Thresher RR, Acierno JS, Jr., Shagoury JK, et al. The GPR54 gene as a regulator of puberty. N Engl J Med. 2003;349(17):1614–27.

5. de Roux N, Genin E, Carel JC, Matsuda F, Chaussain JL, and Milgrom E. Hypogonadotropic hypogonadism due to loss of function of the KiSS1-derived peptide receptor GPR54. Proc Natl Acad Sci U S A. 2003;100(19):10972–6.

6. Spergel DJ. Neuropeptidergic modulation of GnRH neuronal activity and GnRH secretion controlling reproduction: insights from recent mouse studies. Cell Tissue Res. 2019;375(1):179–91.

7. Kim SH. Congenital Hypogonadotropic Hypogonadism and Kallmann Syndrome: Past, Present, and Future. Endocrinol Metab (Seoul). 2015;30(4):456–66.

8. Stamou MI, and Georgopoulos NA. Kallmann syndrome: phenotype and genotype of hypogonadotropic hypogonadism. Metabolism. 2018;86:124–34.

9. Novaira HJ, Sonko ML, Hoffman G, Koo Y, Ko C, Wolfe A, et al. Disrupted kisspeptin signaling in GnRH neurons leads to hypogonadotrophic hypogonadism. Molecular endocrinology. 2014;28(2):225–38.

10. Margolin DH, Kousi M, Chan YM, Lim ET, Schmahmann JD, Hadjivassiliou M, et al. Ataxia, dementia, and hypogonadotropism caused by disordered ubiquitination. N Engl J Med. 2013;368(21):1992–2003.

11. Topaloglu AK, Lomniczi A, Kretzschmar D, Dissen GA, Kotan LD, McArdle CA, et al. Loss-of-function mutations in PNPLA6 encoding neuropathy target esterase underlie pubertal failure and neurological deficits in Gordon Holmes syndrome. The Journal of clinical endocrinology and metabolism. 2014;99(10):E2067–75.

12. Tetreault M, Choquet K, Orcesi S, Tonduti D, Balottin U, Teichmann M, et al. Recessive mutations in POLR3B, encoding the second largest subunit of Pol III, cause a rare hypomyelinating leukodystrophy. American journal of human genetics. 2011;89(5):652–5.

13. Bernard G, Chouery E, Putorti ML, Tetreault M, Takanohashi A, Carosso G, et al. Mutations of POLR3A encoding a catalytic subunit of RNA polymerase Pol III cause a recessive hypomyelinating leukodystrophy. American journal of human genetics. 2011;89(3):415–23.

14. Seminara SB, Acierno JS, Jr., Abdulwahid NA, Crowley WF, Jr., and Margolin DH. Hypogonadotropic hypogonadism and cerebellar ataxia: detailed phenotypic characterization of a large, extended kindred. J Clin Endocrinol Metab. 2002;87(4):1607–12.

15. Abs R, Van Vleymen E, Parizel PM, Van Acker K, Martin M, and Martin JJ. Congenital cerebellar hypoplasia and hypogonadotropic hypogonadism. J Neurol Sci. 1990;98(2-3):259–65.

16. Ueno H, Yamaguchi H, Katakami H, and Matsukura S. A case of Kallmann syndrome associated with Dandy-Walker malformation. Exp Clin Endocrinol Diabetes. 2004;112(1):62–7.

17. Aluclu MU, Bahceci S, and Bahceci M. A rare embryological malformation of brain - Dandy-Walker syndrome - and its association with Kallmann’s syndrome. Neuro Endocrinol Lett. 2007;28(3):255–8.

18. Basson MA, Echevarria D, Ahn CP, Sudarov A, Joyner AL, Mason IJ, et al. Specific regions within the embryonic midbrain and cerebellum require different levels of FGF signaling during development. Development. 2008;135(5):889–98.

19. Yu T, Meiners LC, Danielsen K, Wong MT, Bowler T, Reinberg D, et al. Deregulated FGF and homeotic gene expression underlies cerebellar vermis hypoplasia in CHARGE syndrome. Elife. 2013;2:e01305.

20. Hoshino M, Nakamura S, Mori K, Kawauchi T, Terao M, Nishimura YV, et al. Ptf1a, a bHLH transcriptional gene, defines GABAergic neuronal fates in cerebellum. Neuron. 2005;47(2):201–13.

21. Pascual M, Abasolo I, Mingorance-Le Meur A, Martinez A, Del Rio JA, Wright CV, et al. Cerebellar GABAergic progenitors adopt an external granule cell-like phenotype in the absence of Ptf1a transcription factor expression. Proc Natl Acad Sci U S A. 2007;104(12):5193–8.

22. Ben-Arie N, Bellen HJ, Armstrong DL, McCall AE, Gordadze PR, Guo Q, et al. Math1 is essential for genesis of cerebellar granule neurons. Nature. 1997;390(6656):169–72.

23. Wang VY, Rose MF, and Zoghbi HY. Math1 expression redefines the rhombic lip derivatives and reveals novel lineages within the brainstem and cerebellum. Neuron. 2005;48(1):31–43.

24. Machold R, and Fishell G. Math1 is expressed in temporally discrete pools of cerebellar rhombic-lip neural progenitors. Neuron. 2005;48(1):17–24.

25. Millen KJ, Steshina EY, Iskusnykh IY, and Chizhikov VV. Transformation of the cerebellum into more ventral brainstem fates causes cerebellar agenesis in the absence of Ptf1a function. Proceedings of the National Academy of Sciences of the United States of America. 2014;111(17):E1777–86.

26. Fog CK, Galli GG, and Lund AH. PRDM proteins: important players in differentiation and disease. Bioessays. 2012;34(1):50–60.

27. Fumasoni I, Meani N, Rambaldi D, Scafetta G, Alcalay M, and Ciccarelli FD. Family expansion and gene rearrangements contributed to the functional specialization of PRDM genes in vertebrates. BMC Evol Biol. 2007;7:187.

28. Kim KC, Geng L, and Huang S. Inactivation of a histone methyltransferase by mutations in human cancers. Cancer research. 2003;63(22):7619–23.

29. Hayashi K, Yoshida K, and Matsui Y. A histone H3 methyltransferase controls epigenetic events required for meiotic prophase. Nature. 2005;438(7066):374–8.

30. Di Zazzo E, De Rosa C, Abbondanza C, and Moncharmont B. PRDM Proteins: Molecular Mechanisms in Signal Transduction and Transcriptional Regulation. Biology (Basel). 2013;2(1):107–41.

31. Davis CA, Haberland M, Arnold MA, Sutherland LB, McDonald OG, Richardson JA, et al. PRISM/PRDM6, a transcriptional repressor that promotes the proliferative gene program in smooth muscle cells. Mol Cell Biol. 2006;26(7):2626–36.

32. Ancelin K, Lange UC, Hajkova P, Schneider R, Bannister AJ, Kouzarides T, et al. Blimp1 associates with Prmt5 and directs histone arginine methylation in mouse germ cells. Nat Cell Biol. 2006;8(6):623–30.

33. Gyory I, Wu J, Fejer G, Seto E, and Wright KL. PRDI-BF1 recruits the histone H3 methyltransferase G9a in transcriptional silencing. Nature immunology. 2004;5(3):299–308.

34. Hohenauer T, and Moore AW. The Prdm family: expanding roles in stem cells and development. Development. 2012;139(13):2267–82.

35. Watanabe S, Sanuki R, Sugita Y, Imai W, Yamazaki R, Kozuka T, et al. Prdm13 regulates subtype specification of retinal amacrine interneurons and modulates visual sensitivity. J Neurosci. 2015;35(20):8004–20.

36. Goodson NB, Nahreini J, Randazzo G, Uruena A, Johnson JE, and Brzezinski JAt. Prdm13 is required for Ebf3+ amacrine cell formation in the retina. Dev Biol. 2018;434(1):149–63.

37. Mona B, Uruena A, Kollipara RK, Ma Z, Borromeo MD, Chang JC, et al. Repression by PRDM13 is critical for generating precision in neuronal identity. Elife. 2017;6.

38. Chang JC, Meredith DM, Mayer PR, Borromeo MD, Lai HC, Ou YH, et al. Prdm13 mediates the balance of inhibitory and excitatory neurons in somatosensory circuits. Dev Cell. 2013;25(2):182–95.

39. Hoveyda N, Shield JP, Garrett C, Chong WK, Beardsall K, Bentsi-Enchill E, et al. Neonatal diabetes mellitus and cerebellar hypoplasia/agenesis: report of a new recessive syndrome. J Med Genet. 1999;36(9):700–4.

40. Fujiyama T, Miyashita S, Tsuneoka Y, Kanemaru K, Kakizaki M, Kanno S, et al. Forebrain Ptf1a Is Required for Sexual Differentiation of the Brain. Cell Rep. 2018;24(1):79–94.

41. Segal TY, Mehta A, Anazodo A, Hindmarsh PC, and Dattani MT. Role of gonadotropin-releasing hormone and human chorionic gonadotropin stimulation tests in differentiating patients with hypogonadotropic hypogonadism from those with constitutional delay of growth and puberty. J Clin Endocrinol Metab. 2009;94(3):780–5.

42. Schwarz JM, Cooper DN, Schuelke M, and Seelow D. MutationTaster2: mutation prediction for the deep-sequencing age. Nat Methods. 2014;11(4):361–2.

43. Sun XJ, Xu PF, Zhou T, Hu M, Fu CT, Zhang Y, et al. Genome-wide survey and developmental expression mapping of zebrafish SET domain-containing genes. PLoS One. 2008;3(1):e1499.

44. Satoh A, Brace CS, Rensing N, and Imai S. Deficiency of Prdm13, a dorsomedial hypothalamus-enriched gene, mimics age-associated changes in sleep quality and adiposity. Aging Cell. 2015;14(2):209–18.

45. Spergel DJ. Modulation of Gonadotropin-Releasing Hormone Neuron Activity and Secretion in Mice by Non-peptide Neurotransmitters, Gasotransmitters, and Gliotransmitters. Front Endocrinol (Lausanne). 2019;10:329.

46. Suyama S, and Yada T. New insight into GABAergic neurons in the hypothalamic feeding regulation. J Physiol Sci. 2018;68(6):717–22.

47. Cravo RM, Margatho LO, Osborne-Lawrence S, Donato J, Jr., Atkin S, Bookout AL, et al. Characterization of Kiss1 neurons using transgenic mouse models. Neuroscience. 2011;173:37–56.

48. Prevot V. Knobil and Neill’s Physiology of Reproduction Elsevier; 2015.

49. Knoll JG, Clay CM, Bouma GJ, Henion TR, Schwarting GA, Millar RP, et al. Developmental profile and sexually dimorphic expression of kiss1 and kiss1r in the fetal mouse brain. Frontiers in endocrinology. 2013;4:140.

50. Lapatto R, Pallais JC, Zhang D, Chan YM, Mahan A, Cerrato F, et al. Kiss1-/-mice exhibit more variable hypogonadism than Gpr54-/-mice. Endocrinology. 2007;148(10):4927–36.

51. Barrionuevo F, Georg I, Scherthan H, Lecureuil C, Guillou F, Wegner M, et al. Testis cord differentiation after the sex determination stage is independent of Sox9 but fails in the combined absence of Sox9 and Sox8. Dev Biol. 2009;327(2):301–12.

52. Brown AM, Arancillo M, Lin T, Catt DR, Zhou J, Lackey EP, et al. Molecular layer interneurons shape the spike activity of cerebellar Purkinje cells. Sci Rep. 2019;9(1):1742.

53. Maricich SM, and Herrup K. Pax-2 expression defines a subset of GABAergic interneurons and their precursors in the developing murine cerebellum. J Neurobiol. 1999;41(2):281–94.

54. Zordan P, Croci L, Hawkes R, and Consalez GG. Comparative analysis of proneural gene expression in the embryonic cerebellum. Dev Dyn. 2008;237(6):1726–35.

55. Wulff P, Schonewille M, Renzi M, Viltono L, Sassoe-Pognetto M, Badura A, et al. Synaptic inhibition of Purkinje cells mediates consolidation of vestibulo-cerebellar motor learning. Nat Neurosci. 2009;12(8):1042–9.

56. Heiney SA, Kim J, Augustine GJ, and Medina JF. Precise control of movement kinematics by optogenetic inhibition of Purkinje cell activity. J Neurosci. 2014;34(6):2321–30.

57. Jelitai M, Puggioni P, Ishikawa T, Rinaldi A, and Duguid I. Dendritic excitation-inhibition balance shapes cerebellar output during motor behaviour. Nat Commun. 2016;7:13722.

58. Santens P, Van Damme T, Steyaert W, Willaert A, Sablonniere B, De Paepe A, et al. RNF216 mutations as a novel cause of autosomal recessive Huntington-like disorder. Neurology. 2015;84(17):1760–6.

59. Alqwaifly M, and Bohlega S. Ataxia and Hypogonadotropic Hypogonadism with Intrafamilial Variability Caused by RNF216 Mutation. Neurol Int. 2016;8(2):6444.

60. Calandra CR, Mocarbel Y, Vishnopolska SA, Toneguzzo V, Oliveri J, Cazado EC, et al. Gordon Holmes Syndrome Caused by RNF216 Novel Mutation in 2 Argentinean Siblings. Mov Disord Clin Pract. 2019;6(3):259–62.

61. Shi CH, Schisler JC, Rubel CE, Tan S, Song B, McDonough H, et al. Ataxia and hypogonadism caused by the loss of ubiquitin ligase activity of the U box protein CHIP. Hum Mol Genet. 2014;23(4):1013–24.

62. Sonmez S, Forsyth RJ, Matthews DS, Clarke M, and Splitt M. Oliver-McFarlane syndrome (chorioretinopathy-pituitary dysfunction) with prominent early pituitary dysfunction: differentiation from choroideremia-hypopituitarism. Clin Dysmorphol. 2008;17(4):265–7.

63. Hufnagel RB, Arno G, Hein ND, Hersheson J, Prasad M, Anderson Y, et al. Neuropathy target esterase impairments cause Oliver-McFarlane and Laurence-Moon syndromes. J Med Genet. 2015;52(2):85–94.

64. Synofzik M, Gonzalez MA, Lourenco CM, Coutelier M, Haack TB, Rebelo A, et al. PNPLA6 mutations cause Boucher-Neuhauser and Gordon Holmes syndromes as part of a broad neurodegenerative spectrum. Brain. 2014;137(Pt 1):69–77.

65. Biehl MJ, and Raetzman LT. Rbpj-kappa mediated Notch signaling plays a critical role in development of hypothalamic Kisspeptin neurons. Developmental biology. 2015;406(2):235–46.

66. Oleari R, Lettieri A, Paganoni A, Zanieri L, and Cariboni A. Semaphorin Signaling in GnRH Neurons: From Development to Disease. Neuroendocrinology. 2019;109(3):193–9.

67. Popa SM, Moriyama RM, Caligioni CS, Yang JJ, Cho CM, Concepcion TL, et al. Redundancy in Kiss1 expression safeguards reproduction in the mouse. Endocrinology. 2013;154(8):2784–94.

68. Wang SS, Kloth AD, and Badura A. The cerebellum, sensitive periods, and autism. Neuron. 2014;83(3):518–32.

69. Andreasen NC, and Pierson R. The role of the cerebellum in schizophrenia. Biological psychiatry. 2008;64(2):81–8.

70. Stoodley CJ. The Cerebellum and Neurodevelopmental Disorders. Cerebellum. 2016;15(1):34–7.

71. Gao R, and Penzes P. Common mechanisms of excitatory and inhibitory imbalance in schizophrenia and autism spectrum disorders. Curr Mol Med. 2015;15(2):146–67.

72. McKenna A, Hanna M, Banks E, Sivachenko A, Cibulskis K, Kernytsky A, et al. The Genome Analysis Toolkit: a MapReduce framework for analyzing next-generation DNA sequencing data. Genome Res. 2010;20(9):1297–303.

73. DePristo MA, Banks E, Poplin R, Garimella KV, Maguire JR, Hartl C, et al. A framework for variation discovery and genotyping using next-generation DNA sequencing data. Nat Genet. 2011;43(5):491–8.

74. Whittaker DE, Riegman KL, Kasah S, Mohan C, Yu T, Sala BP, et al. The chromatin remodeling factor CHD7 controls cerebellar development by regulating reelin expression. J Clin Invest. 2017;127(3):874–87.

75. Cariboni A, Davidson K, Rakic S, Maggi R, Parnavelas JG, and Ruhrberg C. Defective gonadotropin-releasing hormone neuron migration in mice lacking SEMA3A signalling through NRP1 and NRP2: implications for the aetiology of hypogonadotropic hypogonadism. Hum Mol Genet. 2011;20(2):336–44.

76. Yaguchi Y, Yu T, Ahmed MU, Berry M, Mason I, and Basson MA. Fibroblast growth factor (FGF) gene expression in the developing cerebellum suggests multiple roles for FGF signaling during cerebellar morphogenesis and development. Dev Dyn. 2009;238(8):2058–72.

77. Gregory LC. Investigation of new candidate genes in a cohort of patients with familial congenital hypopituitarism and associated disorders. Doctoral thesis, URI: 1541141. London: University College London; 2017.

78. Cariboni A, Andre V, Chauvet S, Cassatella D, Davidson K, Caramello A, et al. Dysfunctional SEMA3E signaling underlies gonadotropin-releasing hormone neuron deficiency in Kallmann syndrome. J Clin Invest. 2015;125(6):2413–28.

79. Oleari R, Caramello A, Campinoti S, Lettieri A, Ioannou E, Paganoni A, et al. PLXNA1 and PLXNA3 cooperate to pattern the nasal axons that guide gonadotropin-releasing hormone neurons. Development. 2019;146(21).

80. Bachmann SO, Sledziowska M, Cross E, Kalbassi S, Waldron S, Chen F, et al. Behavioral training rescues motor deficits in Cyfip1 haploinsufficiency mouse model of autism spectrum disorders. Transl Psychiatry. 2019;9(1):29.

81. Howard SR, Oleari R, Poliandri A, Chantzara V, Fantin A, Ruiz-Babot G, et al. HS6ST1 Insufficiency Causes Self-Limited Delayed Puberty in Contrast With Other GnRH Deficiency Genes. J Clin Endocrinol Metab. 2018;103(9):3420–9.

