## Supplemental information for "A recessive *PRDM13* mutation results in congenital hypogonadotropic hypogonadism and cerebellar hypoplasia"

### Supplemental Table 1: Summary of patient phenotypes

| Patient | Age range at Testing (years) | Karyotype | Birth weight Kg (SDS) | GnRH test (IU/L) | Time (mins) | | | hCG test: Serum Free Testosterone (nmol/L) | Scan: MRI or CT | Other features | Treatment (age range started in years) |
|---|---|---|---|---|---|---|---|---|---|---|---|
| | | | | | 0 | +20 | +60 | | | | |
| 1 | 10-15 | XY | 2.8<br>(-1.85) | LH | 0.1 | 2.3 | 2.3 | Baseline: 2.3<br><br>After 3 day hCG (day 4): 2.2 | MRI:<br>- cerebellar hypoplasia - hemispheres and vermis | Delayed motor development<br>BL orchidopexy, Repeated left-side<br>Right-sided thoracic progressive scoliosis: 2-stage surgical fixation of the spine<br>Hypotonia<br>Symmetrical hypo-reflexia<br>Ataxic gait<br>Moderate intellectual deficit | Testosterone (10-15) |
| | | | | FSH | 0.2 | 2.6 | 4.4 | | | | |
| 2 | 10-15 | XX | 3.4<br>(-0.11) | LH | <0.1 | 2.4 | 2.6 | N/A | MRI: Normal | Generalised hypotonia<br>Hyporeflexia<br>Delayed gross motor development<br>Right-sided thoraco-lumbar progressive scoliosis: surgical fixation of the spine<br>Moderate intellectual deficit | Ethinyloestradiol (10-15)<br>Norethisterone (10-15) |
| | | | | FSH | 0.8 | 3.1 | 6.4 | | | | |
| 3 | 10-15 | XY | 2.95<br>(-1.47) | LH | <0.1 | 1.0 | 0.9 | Baseline: <0.69<br><br>After 3 day hCG (day 4): 2.2<br><br>After 3 weeks hCG (day 20): 30.7 | CT: cerebellar hypoplasia | Global developmental delay<br>Generalised hypertonia and hyper-reflexia<br>Strabismus<br>Scoliosis: corrective surgery<br>Wheelchair-dependent<br>BL orchidopexy<br>Severe intellectual deficit<br>Epilepsy | Testosterone (10-15)<br>Lamotrigine (15-20) |
| | | | | FSH | 0.4 | 1.7 | 3.1 | | | | |

**Patients clinical data:**

### Pedigree I

Patient 1 is a patient born to a consanguineous union with a birth weight of 2.8 kg (-1.85 SDS). He was diagnosed with delayed motor development between 0-5 years of age, and noted to have a right-sided thoracic scoliosis. Neurological examination (0-5 years) revealed hypotonia, symmetrical hypo-reflexia, and an ataxic gait. He had bilateral undescended testes and underwent bilateral orchidopexy, which was repeated on the left 3 years later. CT scan on the brain revealed hypoplasia of the cerebellar hemispheres and vermis (Figure 1B). Progression of scoliosis necessitated a 2-stage surgical fixation of the spine at 10-15 years of age. He was referred to the Paediatric Endocrine Clinic 4 years later with delayed puberty. Pubertal staging at this age was G1 P1 A1 -/03 mL, and his height was 136 cm (-3.22 SDS). He had a healthy 18 year old sister, and a 2 year old relative with neurological problems who was eventually diagnosed to have HH (Patient 2). A gonadotropin-releasing hormone (GnRH) test revealed a peak LH of 2.3 IU/L, with an FSH of 4.4 IU/L. A 3-day hCG test revealed no change in the testosterone concentration after 3 hCG injections (peak testosterone of 2.2 nmol/L), and was therefore suboptimal and consistent with hypogonadotrophic hypogonadism (Segal TY et al J Clin Endocrinol Metab. 2009 Mar;94(3):780-5). All other pituitary function tests (including thyroid function tests) were normal. Treatment with exogenous intramuscular testosterone (testosterone esters) was commenced at 10-15 years of age with a gradual increase in dosage over 2 years. Treatment was switched to a transdermal patch preparation at 15-20 years of age, and to long-acting testosterone undecanoate, by intramuscular injection every 3 months, at 20-25 years. He now has moderate intellectual deficit, and remains on testosterone replacement therapy.

Patient 2, sister relative of Patient 1, had a birth weight of 3.4 kg (-0.11 SDS). At postnatal follow-up, she was noted to have generalised hypotonia and hyporeflexia, as well as delayed gross motor development. A progressive right-sided thoraco-lumbar scoliosis was first noted at a similar age to Patient 1. All neurological investigations, including metabolic screen, EMG, and brain MRI were reported as normal. She was first seen in the Paediatric Endocrinology Clinic at the 10-15 years of age, with a similar neurological condition Patient 1. She had not entered spontaneous puberty by the age of 12.5 years, with low basal gonadotrophins (basal LH <0.1 U/L, FSH 0.8 IU/L), and with a peak LH of 2.6 IU/L and an FSH of 6.4 IU/L on GnRH testing.

All other pituitary function tests (including thyroid function and serum prolactin) were normal. When her pubertal staging remained at Tanner B1, she was started on oral ethinyloestradiol to induce pubertal development. The dose of ethinyloestradiol was increased gradually over the subsequent 2 years, which resulted in normal breast development (Tanner B4 by 15 years of age). Oral norethisterone was introduced shortly after in order to induce regular menstrual periods. Surgical fixation of the spine was recently performed at 15-20 years of age. She now has moderate intellectual deficit, and remains on oral ethinyloestradiol and norethisterone.

### Pedigree II

Patient 3 was born to healthy, non-consanguineous parents, with a birth weight of 2.95 kg (-1.47 SDS). The pregnancy was complicated by hyperemesis gravidarum and polyhydramnios, with reported intrauterine growth restriction. Global developmental delay, generalised hypertonia and hyper-reflexia were first noted at 3 months of age. He also had bilateral epicanthic folds and downturned angles of the mouth. A CT brain scan revealed cerebellar hypoplasia. Initially, he was able to walk with a very broad-based gait using a walking frame with significant support. He needed corrective surgery for strabismus as well as spinal surgery for progressive scoliosis at 10-15 years of age, but became completely wheelchair-dependent a year later. He was referred to the Paediatric Endocrine Clinic at 10-15 years of age with a micropenis. On pubertal staging at this age, his stretched penile length was 4 cm (less than P10) and both testes were impalpable. Basal gonadotrophins were low (LH <0.1 IU/L, FSH 0.4 IU/L). A GnRH test performed at 10-15 years of age revealed a peak LH of 0.9 IU/L with an FSH of 3.1 IU/L. The peak testosterone was sub-optimal at 2.3 nmol/L after a 3 day hCG test, with an excellent peak of 30.7 nmol/L after 3 weeks of HCG. Following these tests, the left testis descended into the scrotum (2 mL volume), but the right testis remained impalpable. Exogenous testosterone (testosterone enantate) was commenced at low dose by intramuscular injection at 10-15 years of age, and the dose increased gradually over the following 2 years. Bilateral orchidopexies were performed shortly after. Over time, he experienced penile growth, but both testes remained 2 mL in volume. He currently has severe intellectual deficit, and continues to receive exogenous testosterone replacement therapy. He also began treatment with oral lamotrigine for epilepsy at 10-15 years of age. He has a healthy brother who is 2 years older, and who had gone through puberty spontaneously at the expected age.

### Supplemental Table 2: Neurological phenotype of affected patients

| Information | | Patient 1 | Patient 2 | Patient 3 (Pedigree 2) |
|---|---|---|---|---|
| Neurological information | Tone | Generalised hypotonia | Generalised hypotonia | Generalised hypotonia |
| | Power | Reduced | Very reduced | Extremely reduced |
| | Reflexes | Symmetrical hyporeflexia<br>Down-going plantar reflexes | Symmetrical hyporeflexia<br>Down-going plantar reflexes | Symmetrical hyporeflexia<br>Down-going plantar reflexes |
| | Gait | Broad-based, but able to ambulate by using a frame | Broad-based, but able to ambulate by using a frame | Initially very broad-based, later completely wheelchair-dependent |
| | Nystagmus | Not present | Not present | Present |
| | Intention tremor | Present | Present | Present |
| | Dysdiadochokinesis | Present | Present | Present |
| | Past-pointing | Present | Present | Present |
| Age measured | | 28 years | 17 years | 17 years |
| Weight | | 62 kg | 56 kg | 36.4 kg |
| Height | | 155.4 cm | 144.5 cm | Impossible |
| BMI | | 25.6 $kg/m^2$ | 26.8 $kg/m^2$ | ? |

**Supplemental Table 3: A region on chromosome 6 surrounding the *PRDM13* mutation (c.398-3_407delCAGGGGAGGAGCG), showing the long-affected only haplotype (chr6:98896667-chr6:100480906=1584239bp/1.6Mb) and the nested-shared haplotype (chr6:100053626-chr6:100265121=211495bp/0.2Mb). Genotype data was generated from Illumina Infinium OmniExpress-48 microarray, all genomic positions correspond to genome build GRCh37/Hg19.**

| | | | Patient 1 | Patient 2 | Patient 3 | Control | |
|---|---|---|---|---|---|---|---|
| Name | Chr | Position | Hom | Hom | Hom | Het Carrier | |
| rs3104095 | 6 | 98883438 | AA | AA | AB | BB | |
| rs1494777 | 6 | 98892120 | AA | AA | AB | AB | |
| rs3123339 | 6 | 98893182 | BB | BB | AB | AB | |
| rs3125575 | 6 | 98895069 | AA | AA | AB | AB | |
| rs9321063 | 6 | 98896667 | AA | AA | AA | AA | Beginning of long-affected only haplotype |
| rs17761570 | 6 | 98904572 | AB | AB | AB | AB | |
| rs9388482 | 6 | 98905933 | BB | BB | BB | AB | |
| rs9491633 | 6 | 98914680 | BB | BB | BB | BB | |
| rs9491646 | 6 | 98927363 | BB | BB | BB | BB | |
| rs210400 | 6 | 98931278 | AA | AA | AA | AA | |
| rs169750 | 6 | 98931716 | BB | BB | BB | BB | |
| rs9398813 | 6 | 98935310 | BB | BB | BB | BB | |
| rs9321067 | 6 | 98936578 | BB | BB | BB | BB | |
| rs17058578 | 6 | 98938023 | BB | BB | BB | BB | |
| rs9401901 | 6 | 98938237 | AA | AA | AA | AB | |
| rs9388508 | 6 | 98938249 | BB | BB | BB | BB | |
| rs2227121 | 6 | 98938834 | AB | AB | BB | AB | |
| rs158773 | 6 | 98949550 | BB | BB | BB | BB | |
| rs158774 | 6 | 98953633 | AA | AA | AA | AB | |
| rs158777 | 6 | 98959605 | AA | AA | AA | AB | |
| rs150396 | 6 | 98962591 | BB | BB | BB | BB | |
| rs1481449 | 6 | 98971740 | BB | BB | BB | BB | |

| | | | | | | |
|---|---|---|---|---|---|---|
| rs211215 | 6 | 98975810 | AA | AA | AA | AA |
| rs11961327 | 6 | 98978496 | BB | BB | BB | BB |
| rs211221 | 6 | 98980113 | BB | BB | BB | BB |
| rs211222 | 6 | 98980147 | BB | BB | BB | AB |
| rs17058667 | 6 | 98991464 | BB | BB | BB | BB |
| rs183316 | 6 | 98992369 | BB | BB | BB | BB |
| rs211227 | 6 | 99001499 | AA | AA | AA | AA |
| rs2046174 | 6 | 99013740 | AA | AA | AA | AA |
| rs17829357 | 6 | 99017732 | BB | BB | BB | BB |
| rs9491794 | 6 | 99019431 | BB | BB | BB | AB |
| rs9401968 | 6 | 99027074 | BB | BB | BB | BB |
| rs1481440 | 6 | 99032723 | AA | AA | AA | AB |
| rs11752460 | 6 | 99033489 | AA | AA | AA | AA |
| rs1481438 | 6 | 99034313 | AA | AA | AA | AB |
| rs6929790 | 6 | 99045749 | AA | AA | AA | AB |
| rs17830067 | 6 | 99050825 | BB | BB | BB | BB |
| rs969540 | 6 | 99058170 | BB | BB | BB | BB |
| rs17058737 | 6 | 99076746 | AA | AA | AA | AA |
| rs11757142 | 6 | 99080658 | AA | AA | AA | AA |
| rs17058761 | 6 | 99092539 | BB | BB | BB | BB |
| rs11752997 | 6 | 99124184 | AA | AA | AA | AA |
| rs4472356 | 6 | 99126082 | AA | AA | AA | AB |
| rs9375573 | 6 | 99126994 | AA | AA | AA | AB |
| rs12204275 | 6 | 99138875 | BB | BB | BB | BB |
| rs6939572 | 6 | 99139350 | AA | AA | AA | AA |
| rs6924761 | 6 | 99140492 | BB | BB | BB | BB |
| rs4424090 | 6 | 99147074 | BB | BB | BB | BB |
| rs9375604 | 6 | 99170039 | AA | AA | AA | AA |
| rs4529305 | 6 | 99173425 | AA | AA | AA | AA |
| rs9388676 | 6 | 99175625 | BB | BB | BB | BB |
| rs11154467 | 6 | 99195548 | BB | BB | BB | AB |

| | | | | | | |
|---|---|---|---|---|---|---|
| rs4406241 | 6 | 99203222 | BB | BB | BB | BB |
| rs7762570 | 6 | 99214884 | AA | AA | AA | AB |
| rs4839976 | 6 | 99218958 | AA | AA | AA | AB |
| rs4398748 | 6 | 99219522 | AA | AA | AA | AA |
| rs9385493 | 6 | 99221669 | AA | AA | AA | AB |
| rs10499023 | 6 | 99226729 | AA | AA | AA | AA |
| rs4839977 | 6 | 99229574 | BB | BB | BB | AB |
| rs4555920 | 6 | 99230880 | BB | BB | BB | AB |
| rs3923049 | 6 | 99233448 | AA | AA | AA | AB |
| rs9375642 | 6 | 99235909 | AA | AA | AA | AB |
| rs9402154 | 6 | 99236649 | BB | BB | BB | AB |
| rs9388719 | 6 | 99237315 | BB | BB | BB | AA |
| rs12174549 | 6 | 99237479 | AA | AA | AA | AA |
| rs7762652 | 6 | 99242592 | BB | BB | BB | AA |
| rs9385517 | 6 | 99250916 | BB | BB | BB | BB |
| rs4839986 | 6 | 99251754 | BB | BB | BB | AB |
| rs6569636 | 6 | 99258445 | AA | AA | AA | BB |
| rs9375688 | 6 | 99270438 | BB | BB | BB | AB |
| rs9375689 | 6 | 99270533 | BB | BB | BB | AA |
| rs2444935 | 6 | 99275290 | BB | BB | BB | BB |
| rs1869641 | 6 | 99277867 | AA | AA | AA | BB |
| rs1883306 | 6 | 99279449 | AA | AA | AA | AB |
| rs3823036 | 6 | 99284532 | AA | AA | AA | AB |
| rs195853 | 6 | 99290334 | BB | BB | BB | BB |
| rs195852 | 6 | 99290592 | AA | AA | AA | BB |
| rs195851 | 6 | 99294322 | BB | BB | BB | AA |
| rs174447 | 6 | 99309057 | AA | AA | AA | BB |
| rs9388789 | 6 | 99320715 | BB | BB | BB | AB |
| rs11537982 | 6 | 99323424 | BB | BB | BB | BB |
| rs9375728 | 6 | 99326490 | AA | AA | AA | AB |
| rs195831 | 6 | 99328730 | AA | AA | AA | AA |

| | | | | | | |
|---|---|---|---|---|---|---|
| rs10484609 | 6 | 99336279 | AA | AA | AA | AB |
| rs7739884 | 6 | 99346145 | BB | BB | BB | AB |
| rs195824 | 6 | 99351047 | AA | AA | AA | AB |
| rs1011676 | 6 | 99374400 | BB | BB | BB | BB |
| rs17058986 | 6 | 99431200 | AA | AA | AA | AA |
| rs196959 | 6 | 99432747 | BB | BB | BB | BB |
| rs196960 | 6 | 99433614 | BB | BB | BB | AA |
| rs2747734 | 6 | 99437566 | BB | BB | BB | AA |
| rs2572109 | 6 | 99440369 | BB | BB | BB | AB |
| rs12200990 | 6 | 99441083 | BB | BB | BB | AB |
| rs2180046 | 6 | 99450081 | BB | BB | BB | BB |
| rs2747739 | 6 | 99462075 | BB | BB | BB | AB |
| rs9402354 | 6 | 99462324 | AA | AA | AA | AA |
| rs9375844 | 6 | 99463771 | BB | BB | BB | BB |
| rs2207446 | 6 | 99464542 | AA | AA | AA | BB |
| rs12173555 | 6 | 99467661 | BB | BB | BB | AB |
| rs9388950 | 6 | 99468942 | AA | AA | AA | BB |
| rs2092772 | 6 | 99473410 | BB | BB | BB | AB |
| rs12201236 | 6 | 99478174 | BB | BB | BB | AB |
| rs4839737 | 6 | 99478304 | AA | AA | AA | BB |
| rs2747748 | 6 | 99479842 | AA | AA | AA | AA |
| rs7756447 | 6 | 99480633 | BB | BB | BB | BB |
| rs11756151 | 6 | 99485266 | BB | BB | BB | BB |
| rs1997937 | 6 | 99487230 | BB | BB | BB | AB |
| rs12173832 | 6 | 99498599 | BB | BB | BB | BB |
| rs2144241 | 6 | 99500963 | AA | AA | AA | AA |
| rs6916751 | 6 | 99501709 | AA | AA | AA | AA |
| rs9493282 | 6 | 99504377 | BB | BB | BB | BB |
| rs12660289 | 6 | 99515134 | AA | AA | AA | AB |
| rs2207445 | 6 | 99521031 | BB | BB | BB | BB |
| rs7764372 | 6 | 99524062 | BB | BB | BB | BB |

| | | | | | | |
|---|---|---|---|---|---|---|
| rs9375894 | 6 | 99524281 | AA | AA | AA | AB |
| rs12212298 | 6 | 99531172 | AA | AA | AA | AA |
| rs9385615 | 6 | 99531774 | BB | BB | BB | AB |
| rs2572098 | 6 | 99538132 | AA | AA | AA | AA |
| rs9402444 | 6 | 99539153 | BB | BB | BB | AB |
| rs6902772 | 6 | 99550464 | BB | BB | BB | AB |
| rs6940049 | 6 | 99551169 | AA | AA | AA | AA |
| rs3860236 | 6 | 99551765 | BB | BB | BB | AB |
| rs10457592 | 6 | 99551970 | BB | BB | BB | BB |
| rs12208335 | 6 | 99552868 | AA | AA | AA | AB |
| rs6938641 | 6 | 99557664 | AA | AA | AA | AA |
| rs9373032 | 6 | 99559165 | BB | BB | BB | AB |
| rs1081025 | 6 | 99559773 | AA | AA | AA | AB |
| rs12526079 | 6 | 99565215 | BB | BB | BB | BB |
| rs7769941 | 6 | 99569709 | AA | AA | AA | AA |
| rs2388839 | 6 | 99574363 | BB | BB | BB | AB |
| rs12111251 | 6 | 99575661 | BB | BB | BB | BB |
| rs4454147 | 6 | 99575874 | BB | BB | BB | BB |
| rs2029964 | 6 | 99583773 | AA | AA | AA | AB |
| rs728758 | 6 | 99584658 | BB | BB | BB | BB |
| rs11154718 | 6 | 99592404 | BB | BB | BB | AB |
| rs12190591 | 6 | 99593258 | AA | AA | AA | AA |
| rs9321394 | 6 | 99600083 | AA | AA | AA | AB |
| rs11754157 | 6 | 99607499 | BB | BB | BB | BB |
| rs4839999 | 6 | 99610074 | BB | BB | BB | AB |
| rs12206927 | 6 | 99613136 | BB | BB | BB | AA |
| rs12110525 | 6 | 99629252 | BB | BB | BB | BB |
| rs9373057 | 6 | 99630038 | BB | BB | BB | AB |
| rs9399068 | 6 | 99633011 | AA | AA | AA | BB |
| rs9375977 | 6 | 99637123 | BB | BB | BB | AA |
| rs7745052 | 6 | 99640610 | BB | BB | BB | AB |

| | | | | | | |
|---|---|---|---|---|---|---|
| rs1496971 | 6 | 99641248 | AA | AA | AA | BB |
| rs1908804 | 6 | 99646240 | AA | AA | AA | AA |
| rs7767885 | 6 | 99659802 | AA | AA | AA | BB |
| rs7769752 | 6 | 99662681 | AA | AA | AA | BB |
| rs17059246 | 6 | 99662743 | AA | AA | AA | AB |
| rs9389116 | 6 | 99664294 | BB | BB | BB | AB |
| rs4458696 | 6 | 99665178 | BB | BB | BB | BB |
| rs9373072 | 6 | 99669676 | BB | BB | BB | AB |
| rs2132683 | 6 | 99672014 | AA | AA | AA | AB |
| rs9375997 | 6 | 99673152 | BB | BB | BB | AB |
| rs9402556 | 6 | 99673466 | AA | AA | AA | BB |
| rs12528619 | 6 | 99675908 | BB | BB | BB | BB |
| rs1566116 | 6 | 99678424 | AA | AA | AA | AA |
| rs4840017 | 6 | 99679578 | BB | BB | BB | BB |
| rs9402564 | 6 | 99679902 | BB | BB | BB | BB |
| rs10155713 | 6 | 99683181 | BB | BB | BB | BB |
| rs2029965 | 6 | 99683802 | BB | BB | BB | BB |
| rs9376014 | 6 | 99690090 | AA | AA | AA | AA |
| rs12193060 | 6 | 99690449 | AA | AA | AA | AA |
| rs1874538 | 6 | 99694494 | AA | AA | AA | AA |
| rs6904604 | 6 | 99695108 | BB | BB | BB | BB |
| rs9493928 | 6 | 99711169 | BB | BB | BB | BB |
| rs9483707 | 6 | 99717329 | BB | BB | BB | AB |
| rs13219146 | 6 | 99717914 | AA | AA | AA | AB |
| rs1045728 | 6 | 99721049 | BB | BB | BB | AB |
| rs12660321 | 6 | 99727653 | BB | BB | BB | BB |
| rs1496979 | 6 | 99728442 | AA | AA | AA | AB |
| rs6933093 | 6 | 99729901 | BB | BB | BB | AB |
| rs9373105 | 6 | 99733939 | AA | AA | AA | AA |
| rs1496980 | 6 | 99734185 | BB | BB | BB | AB |
| rs12207550 | 6 | 99740492 | BB | BB | BB | BB |

| | | | | | | |
|---|---|---|---|---|---|---|
| rs221582 | 6 | 99744107 | BB | BB | BB | AB |
| rs221578 | 6 | 99747633 | AA | AA | AA | AA |
| rs221530 | 6 | 99753499 | AA | AA | AA | AA |
| rs6913076 | 6 | 99763266 | BB | BB | BB | BB |
| rs182613 | 6 | 99768894 | BB | BB | BB | AB |
| rs221527 | 6 | 99771540 | BB | BB | BB | BB |
| rs13206094 | 6 | 99772374 | AA | AA | AA | AB |
| rs4840031 | 6 | 99773417 | BB | BB | BB | BB |
| rs6922449 | 6 | 99776986 | AA | AA | AA | AB |
| rs11757364 | 6 | 99781218 | BB | BB | BB | AB |
| rs17059400 | 6 | 99782879 | AA | AA | AA | AA |
| rs11963108 | 6 | 99793615 | BB | BB | BB | BB |
| rs9402701 | 6 | 99801698 | BB | BB | BB | AB |
| rs12198238 | 6 | 99803269 | BB | BB | BB | AB |
| rs13194648 | 6 | 99804157 | BB | BB | BB | AB |
| rs17059457 | 6 | 99806408 | AA | AA | AA | AA |
| rs4839747 | 6 | 99814530 | BB | BB | BB | BB |
| rs6925344 | 6 | 99819379 | BB | BB | BB | AB |
| rs12193590 | 6 | 99826265 | BB | BB | BB | AB |
| rs9402716 | 6 | 99827314 | BB | BB | BB | AB |
| rs4840038 | 6 | 99828941 | AA | AA | AA | AB |
| rs11154812 | 6 | 99836954 | BB | BB | BB | BB |
| rs9376137 | 6 | 99842056 | BB | BB | BB | BB |
| rs9376138 | 6 | 99842138 | BB | BB | BB | BB |
| rs11961608 | 6 | 99847168 | AA | AA | AA | AA |
| rs4144165 | 6 | 99847260 | BB | BB | BB | BB |
| rs3811072 | 6 | 99851977 | BB | BB | BB | BB |
| rs11154824 | 6 | 99852267 | AA | AA | AA | AA |
| rs4840039 | 6 | 99869689 | BB | BB | BB | BB |
| rs12198321 | 6 | 99871010 | BB | BB | BB | BB |
| rs4351270 | 6 | 99873534 | BB | BB | BB | BB |

| | | | | | | |
|---|---|---|---|---|---|---|
| rs6923983 | 6 | 99877219 | AA | AA | AA | AB |
| rs1134718 | 6 | 99880380 | AA | AA | AA | AA |
| rs12214037 | 6 | 99880572 | AA | AA | AA | AA |
| rs17785525 | 6 | 99882563 | BB | BB | BB | AB |
| rs6570064 | 6 | 99883137 | AA | AA | AA | AB |
| rs9402791 | 6 | 99883694 | AA | AA | AA | AA |
| rs6570065 | 6 | 99883704 | BB | BB | BB | BB |
| rs4839748 | 6 | 99889915 | AA | AA | AA | AB |
| rs9494471 | 6 | 99893527 | BB | BB | BB | AB |
| rs12203426 | 6 | 99893878 | AA | AA | AA | AA |
| rs4504482 | 6 | 99893938 | AA | AA | AA | AA |
| rs10155760 | 6 | 99901608 | BB | BB | BB | BB |
| rs6916603 | 6 | 99905558 | BB | BB | BB | AB |
| rs10457650 | 6 | 99916182 | AA | AA | AA | AA |
| rs7745012 | 6 | 99921822 | BB | BB | BB | BB |
| rs9483935 | 6 | 99933680 | BB | BB | BB | BB |
| rs6918880 | 6 | 99934446 | AA | AA | AA | AA |
| rs12717185 | 6 | 99939045 | AA | AA | AA | AB |
| rs2209157 | 6 | 99964012 | BB | BB | BB | BB |
| rs9402863 | 6 | 99971429 | BB | BB | BB | AB |
| rs10223892 | 6 | 99973893 | BB | BB | BB | AB |
| rs17224695 | 6 | 99974948 | BB | BB | BB | AB |
| rs2057517 | 6 | 99980252 | AA | AA | AA | AA |
| rs2057518 | 6 | 99980522 | AA | AA | AA | AA |
| rs7754710 | 6 | 99986195 | BB | BB | BB | AB |
| rs1054227 | 6 | 99990856 | BB | BB | BB | AB |
| rs2296154 | 6 | 99993268 | AA | AA | AA | AB |
| rs13205324 | 6 | 99995320 | BB | BB | BB | BB |
| rs543967 | 6 | 100005775 | BB | BB | BB | AB |
| rs1590359 | 6 | 100022125 | BB | BB | BB | BB |
| rs514769 | 6 | 100031037 | AA | AA | AA | AB |

| | | | | | | | |
|---|---|---|---|---|---|---|---|
| rs1552855 | 6 | 100046845 | AA | AA | AA | AB | |
| rs9484083 | 6 | 100046933 | AA | AA | AA | AB | |
| rs7741279 | 6 | 100053626 | BB | BB | BB | BB | Beginning of nested-shared haplotype |
| rs330843 | 6 | 100060761 | BB | BB | BB | BB | |
| | **6** | **100060906** | | | | | **Mutation position** |
| rs3734346 | 6 | 100062766 | AA | AA | AA | AA | |
| rs330844 | 6 | 100063243 | BB | BB | BB | BB | |
| rs6927488 | 6 | 100081441 | BB | BB | BB | BB | |
| rs594231 | 6 | 100082983 | BB | BB | BB | BB | |
| rs9402954 | 6 | 100088902 | AA | AA | AA | AA | |
| rs1339203 | 6 | 100092390 | BB | BB | BB | BB | |
| rs546567 | 6 | 100099703 | BB | BB | BB | BB | |
| rs13193313 | 6 | 100108500 | AA | AA | AA | AA | |
| rs9321659 | 6 | 100116092 | BB | BB | BB | BB | |
| rs650783 | 6 | 100125119 | AA | AA | AA | AA | |
| rs9373217 | 6 | 100129570 | BB | BB | BB | BB | |
| rs503649 | 6 | 100129959 | AA | AA | AA | AA | |
| rs472977 | 6 | 100130991 | BB | BB | BB | BB | |
| rs9495145 | 6 | 100131219 | BB | BB | BB | BB | |
| rs9376355 | 6 | 100132920 | AA | AA | AA | AA | |
| rs12661094 | 6 | 100161035 | BB | BB | BB | BB | |
| rs12211649 | 6 | 100172277 | BB | BB | BB | BB | |
| rs4839755 | 6 | 100173173 | BB | BB | BB | BB | |
| rs9389645 | 6 | 100173832 | AA | AA | AA | AA | |
| rs6916754 | 6 | 100174464 | BB | BB | BB | BB | |
| rs6908196 | 6 | 100185261 | BB | BB | BB | BB | |
| rs12527523 | 6 | 100187748 | BB | BB | BB | BB | |
| rs12525414 | 6 | 100192215 | AA | AA | AA | AA | |
| rs17826560 | 6 | 100195578 | AA | AA | AA | AA | |
| rs7742890 | 6 | 100197285 | AA | AA | AA | NC | |
| rs7356874 | 6 | 100197712 | BB | BB | BB | BB | |

| | | | | | | | |
|---|---|---|---|---|---|---|---|
| rs10485226 | 6 | 100206681 | AA | AA | AA | AA | |
| rs10485227 | 6 | 100207327 | BB | BB | BB | BB | |
| rs4388294 | 6 | 100207838 | AA | AA | AA | AA | |
| rs17059765 | 6 | 100208624 | AA | AA | AA | AA | |
| rs4431442 | 6 | 100213515 | BB | BB | BB | BB | |
| rs9403091 | 6 | 100228256 | BB | BB | BB | AB | |
| rs9376435 | 6 | 100231618 | BB | BB | BB | BB | |
| rs9385869 | 6 | 100233278 | AA | AA | AA | AA | |
| rs9399284 | 6 | 100233870 | AA | AA | AA | AA | |
| rs12663112 | 6 | 100236473 | AA | AA | AA | AA | |
| rs4596491 | 6 | 100236866 | BB | BB | BB | BB | |
| rs12198721 | 6 | 100237464 | AA | AA | AA | AA | |
| rs6936455 | 6 | 100237880 | BB | BB | BB | BB | |
| rs9403103 | 6 | 100240267 | BB | BB | BB | BB | |
| rs6929006 | 6 | 100242158 | BB | BB | BB | BB | |
| rs6929428 | 6 | 100242366 | BB | BB | BB | BB | |
| rs17236659 | 6 | 100243033 | AA | AA | AA | AA | |
| rs9376456 | 6 | 100243997 | BB | BB | BB | BB | |
| rs6931227 | 6 | 100247048 | AA | AA | AA | AA | |
| rs7763983 | 6 | 100249106 | BB | BB | BB | BB | |
| rs17059858 | 6 | 100250740 | AA | AA | AA | AA | |
| rs9484314 | 6 | 100251499 | AA | AA | AA | AA | |
| rs17059867 | 6 | 100252886 | BB | BB | BB | BB | |
| rs9373250 | 6 | 100255471 | BB | BB | BB | BB | |
| rs12216503 | 6 | 100257066 | BB | BB | BB | BB | |
| rs4495279 | 6 | 100257668 | BB | BB | BB | BB | |
| rs6934621 | 6 | 100265121 | BB | BB | BB | BB | End of nested-shared haplotype |
| rs7760502 | 6 | 100266123 | AA | AA | AA | AB | |
| rs17059881 | 6 | 100268849 | AA | AA | AA | AA | |
| rs13204333 | 6 | 100277840 | BB | BB | BB | AB | |
| rs9403141 | 6 | 100279956 | BB | BB | BB | BB | |

| | | | | | | |
|---|---|---|---|---|---|---|
| rs9385894 | 6 | 100280032 | BB | BB | BB | BB |
| rs7453413 | 6 | 100281817 | AA | AA | AA | AB |
| rs3922542 | 6 | 100282075 | BB | BB | BB | AB |
| rs9321763 | 6 | 100286127 | BB | BB | BB | BB |
| rs4370372 | 6 | 100288317 | AA | AA | AA | AA |
| rs9389752 | 6 | 100289337 | BB | BB | BB | BB |
| rs9495653 | 6 | 100292091 | AA | AA | AA | AA |
| rs9495657 | 6 | 100293531 | AA | AA | AA | AA |
| rs4990843 | 6 | 100296024 | BB | BB | BB | BB |
| rs9403163 | 6 | 100297481 | AA | AA | AA | AA |
| rs12197810 | 6 | 100317146 | AA | AA | AA | AA |
| rs9403180 | 6 | 100319031 | AA | AA | AA | AA |
| rs2397663 | 6 | 100321669 | AA | AA | AA | AA |
| rs2397664 | 6 | 100321734 | AA | AA | AA | AA |
| rs9403186 | 6 | 100324464 | AA | AA | AA | AA |
| rs9495745 | 6 | 100326214 | BB | BB | BB | BB |
| rs4840097 | 6 | 100328198 | BB | BB | BB | AB |
| rs9389810 | 6 | 100329446 | AA | AA | AA | AB |
| rs2397678 | 6 | 100345238 | AA | AA | AA | AB |
| rs6902801 | 6 | 100345403 | AA | AA | AA | AA |
| rs7751620 | 6 | 100353268 | AA | AA | AA | AB |
| rs9403208 | 6 | 100354935 | BB | BB | BB | BB |
| rs4560657 | 6 | 100366635 | AA | AA | AA | AB |
| rs13206575 | 6 | 100369284 | BB | BB | BB | AB |
| rs4559096 | 6 | 100391872 | BB | BB | BB | AB |
| rs12215494 | 6 | 100392532 | BB | BB | BB | AB |
| rs13195863 | 6 | 100393761 | AA | AA | AA | AB |
| rs10499026 | 6 | 100397526 | AA | AA | AA | AB |
| rs4840106 | 6 | 100403514 | BB | BB | BB | BB |
| rs7758072 | 6 | 100411417 | BB | BB | BB | BB |
| rs7739904 | 6 | 100427015 | BB | BB | BB | BB |

| | | | | | | | |
|---|---|---|---|---|---|---|---|
| rs9496070 | 6 | 100428029 | BB | BB | BB | BB | |
| rs13212643 | 6 | 100428313 | AA | AA | AA | AA | |
| rs12203515 | 6 | 100430062 | BB | BB | BB | BB | |
| rs9496085 | 6 | 100430803 | BB | BB | BB | BB | |
| rs9389934 | 6 | 100431321 | AA | AA | AA | AA | |
| rs2001456 | 6 | 100439009 | BB | BB | BB | AB | |
| rs6925272 | 6 | 100442268 | BB | BB | BB | AB | |
| rs9969034 | 6 | 100442554 | BB | BB | BB | BB | |
| rs2397694 | 6 | 100445429 | AA | AA | AA | AA | |
| rs9399386 | 6 | 100446900 | BB | BB | BB | AB | |
| rs3763374 | 6 | 100448788 | BB | BB | BB | BB | |
| rs9385975 | 6 | 100449301 | BB | BB | BB | BB | |
| rs9389951 | 6 | 100457411 | BB | BB | BB | AB | |
| rs9484578 | 6 | 100469529 | AA | AA | AA | AA | |
| rs4240585 | 6 | 100480906 | BB | BB | BB | AB | End of long-affected only haplotype |
| rs6925134 | 6 | 100482016 | AB | AB | BB | BB | |
| rs12202476 | 6 | 100492824 | BB | BB | BB | BB | |
| rs3890820 | 6 | 100494186 | BB | BB | BB | BB | |
| rs6903499 | 6 | 100510685 | BB | BB | BB | BB | |
| rs11155243 | 6 | 100514432 | BB | BB | BB | BB | |
| rs6938191 | 6 | 100521649 | AB | AB | BB | AB | |

### Primer sequences:

qPCR primer sequences:

| Gene | Forward Primer | Reverse Primer |
|---|---|---|
| ***Prdm13 exon 2-3*** | GGATAGGGTTAATCCGGGCA | TGGAGTCGCAGTTGTAGGGA |
| ***Prdm13 exon 4*** | GAGATCGCCATGCACACACAG | AGTACAGCTTGCCACAGTAGAG |
| ***Kiss1*** | ATGATCTCAATGGCTTCTTGG | CCAGGCATTAACGAGTTCCT |
| ***Npvf*** | CAAGACACCCGCTGATTTGC | TCCTCTCCTCGTTCGCTTTC |
| ***Pomc*** | TGGGCGAGCTGATGACCT | GCCGACTGTGAAATCTGAAAGG |
| ***Agrp*** | CTTTGGCGGAGGTGCTAGAT | AGGACTCGTGCAGCCTTACAC |
| ***Npy*** | TGGCCAGATACTACTCCGCT | TCCTCTCCTCGTTCGCTTTC |
| ***Gad1*** | CTTCTTCAGGCTCTCCCGTG | CAGGAACAGGCTCGGTTCAG |
| ***Gapdh*** | AGGTCGGTGTGAACGGATTTG | TGTAGACCATGTAGTTGAGGTCA |

RT-PCR primer sequences:

| Gene | Forward Primer | Reverse Primer |
|---|---|---|
| ***Prdm13*** | GCCACTTGTGCCTCTACTGT | CCTCCACAGACAAGAGCGTT |
| ***Gapdh*** | TGGCATTGTGGAAGGGCTCATGAC | ATGCCAGTGAGCTTCCCGTTCAGC |

Genotyping primer sequences:

| Gene | Forward Primer | Reverse Primer |
|---|---|---|
| ***Prdm13 - KO*** | CACCTCAGTCTTTGCCTTCCTTGCAA | CTACAACTGCGACTCCAACGCATGAT |
| ***Prdm13 - WT*** | CACCTCAGTCTTTGCCTTCCTTGCAA | CAGAGAAAGAGTACCCTTGTGCCT |
